# Steps against the burden of Parkinson’s disease (StepuP): Protocol of a randomized controlled trial elucidating the biomechanical and neurophysiological mechanisms of a speed dependent treadmill training intervention

**DOI:** 10.64898/2026.03.12.26348248

**Authors:** Anina Moira van Leeuwen, Julius Welzel, Ilaria D’Ascanio, Charlotte Lang, Vaishali Vinod, Pia Görrisen, Johanna Geritz, Clint Hansen, Eran Gazit, Shahar Siman Tov, Revital Prusak, Ilaria Casadei, Angela Contri, Francesca Tampellini, Leonardo Pellicciari, Giovanna Lopane, Giovanna Calandra Buonaura, Luca Palmerini, Noman Zahid, Mayna Ratanapongleka, Husna Razee, Frederic von Wegner, Bernadette C.M. van Wijk, Sjoerd M. Bruijn, Deepak K. Ravi, Yoshiro Okubo, Navrag B. Singh, Matthew Brodie, Fabio La Porta, Jeffrey M. Hausdorff, Walter Maetzler, Jaap H. van Dieën

## Abstract

**Objective:** Parkinson’s disease can impair gait and stability, leading to reduced independence and increased fall risk. While speed dependent treadmill training (SDTT) is clinically effective, the specific biomechanical and neurophysiological mechanisms driving these improvements remain unclear. The “StepuP” multicenter randomized controlled trial aims to elucidate these mechanisms and determine whether training enriched with virtual reality or mechanical perturbations (SDTT+) enhances gait efficacy and transfer to daily life.

**Methods:** We will recruit 126 individuals with Parkinson’s disease across four clinical sites and 21 healthy older adults as a reference group. Participants will be randomized to receive either standard SDTT or SDTT+ for 12 sessions. To capture the trajectory of recovery and retention, assessments will occur at three distinct timepoints: baseline, post-intervention, and a 12-week follow-up, each assessment including synchronized 64-channel electroencephalography (EEG), electromyography (EMG), and 3D kinematics. This multimodal setup allows for the quantification of cortical beta-band activity, corticomuscular coherence, and stability-related foot placement control. Furthermore, we will assess participant’s satisfaction, usability, and engagement through questionnaires and interviews to understand individual adherence and barriers to training.

**Significance:** The primary clinical endpoint is comfortable overground walking speed. We hypothesize that gait improvements are mediated by improved stability-related foot placement and cortical sensorimotor integration. By correlating lab-based mechanistic changes with real-world mobility patterns and participant experiences, this study seeks to identify specific pathophysiological mechanisms engaged during the treadmill training. These insights will help distinguish responders from non-responders, facilitating the development of personalized, acceptable, and effective rehabilitation strategies.

## Introduction

Parkinson’s Disease (PD) is a neurodegenerative disorder that affects approximately 10 million people worldwide, with its prevalence having doubled in the past 25 years [1]. Costs associated with PD are estimated at $52 billion per year in the USA alone [2]. The symptoms of PD can result in a gradual negative impact on an individual’s capacity to perform activities of daily living and to live independently [3,4]. In fact, both disability and mortality might be increasing faster than for any other neurological disorder [5]. While one of the risk factors of PD is age [6], younger adults are also susceptible and age-adjusted models show an increase in PD disease burden, especially in countries with higher socio-demographic indices [5]. If unmitigated, the projected demographic shift towards older adults over the next few decades could result in a large societal, personal and economic burden.

To effectively treat PD, dopaminergic replacement therapy [7] remains the most effective option, particularly for alleviating the cardinal symptoms of bradykinesia, rigidity, and tremor [1]. As the disease progresses, however, these symptoms become more difficult to control with medication, and long-term treatment is often accompanied by complications such as motor fluctuations and dyskinesia [8]. While dopaminergic replacement therapy can provide benefits for gait in the early stages of treatment, gait-related impairments frequently persist or worsen over time, despite medication [8–10] and advanced therapies such as deep brain stimulation (DBS) [9–12]. This is a critical issue, as gait deficits are highly disabling, and up to 70% of individuals with PD experience falls each year [13]. This underscores the need for other forms of therapy to better target gait dysfunction in persons with PD (PwPD).

Accumulating evidence indicates that non-pharmacological approaches including exercise [11,14] can be effective in alleviating cardinal symptoms of PD and may even potentially slow disease progression [12]. Moreover, these treatment approaches have been shown to improve gait quality [15,16] and reduce fall rates [15,17]. Exercise interventions are particularly effective in addressing gait dysfunction in PD [18]. In particular, treadmill training has shown promising results in improving gait performance in PD [16]. This form of training provides externally cued [19,20], repetitive stepping under constant speed conditions, which can increase stride length and walking speed and reduce gait variability [21,22]. However, the underlying biomechanical and neurophysiological mechanisms of this effect are poorly understood.

Improvements in gait performance as evidenced by an increased walking speed and stride length, suggest underlying improvements in anteroposterior foot placement control. From a biomechanical perspective, foot placement control does not only subserve forward progression and speed modulation but is also the most dominant stability control mechanism during (steady-state) walking [23,24]. As a stability mechanism, foot placement is controlled with respect to the position and velocity of the body’s center of mass (CoM) [25]. Such foot placement is realized through feedback control in which the kinematic state of the CoM serves as an important sensory input [28–30]. A linear model, relating the CoM position and velocity to foot placement captures this control mechanism [25]. Fitting this model on treadmill walking data showed that, at certain instances of the swing phase, the CoM state can predict foot placement better than the swing foot itself [25]. Variations in muscle activity [26,27] and sensitivity to sensory perturbations [28] have been associated with the foot placement mechanism [26,27], supporting neural control to maintain steady-state gait stability. Foot placement control is less precise in older compared to younger adults [28] and PwPD show impaired foot placement control compared to age-matched healthy adults [29]. Therefore, it is of interest to assess whether improved gait performance following treadmill training is related to the extent to which stability-related foot placement control is improved.

From a neurophysiological perspective, improved stability-related foot placement control is expected to be achieved through enhanced cortical control. Electroencephalographic (EEG) studies have revealed cortical involvement during walking in the form of gait-related modulations in the beta band [30,31] Although beta modulations during gait are yet to be related to foot placement control, decreased beta power in the left premotor area has been associated with stability control [32]. In PwPD altered beta modulation is considered to maintain an abnormally stable, inflexible motor state [33,34] . An inflexible motor state can impair rapid adjustments during the gait cycle [34], potentially resulting in the observed imprecise foot placement in PD [29]. Improved beta modulation throughout the gait cycle would therefore be expected to manifest alongside potential treadmill training effects on foot placement precision. The role of the cortex in stability-related foot placement control is predicted to be processing multi-modal sensory information of the CoM state [28,35,36] through sensorimotor integration [37], ultimately driving muscle activation underlying foot placement control [38]. Functional connectivity between the cortex and muscles involved in foot placement control can be assessed through corticomuscular coherence [39]. Gait-related beta-band corticomuscular coherence with the tibialis anterior muscle has been shown to be lower in individuals with PD as compared to age-matched people without PD [40]. A comparison of the corticomuscular coherence of muscles specifically related to stability-related foot placement control between age-matched controls and PwPD will allow us to predict the direction of treadmill training effects on this measure of cortical control. Taken together, the biomechanical and neurophysiological perspectives aim at better insights into the mechanisms underlying speed dependent treadmill training.

Insights into the underlying mechanisms might also help us better understand potential transfer of the training effects to daily life. In daily life impaired gait and impaired dynamic stability are even more apparent in PwPD than in a laboratory or clinical setting [41]. The reduced quality of foot placement control [29] might also be reflected in daily life gait, and could lead to greater gait instability and risk of falling. However, it is still uncertain to what extent treadmill training effects translate to daily life [42,43]. In PwPD, only one study has investigated the effectiveness of a multidisciplinary rehabilitation program and found no transfer of improvements to daily life gait [42]. However, individual responses may vary, and sustained adherence to training schemes remains a challenge. Understanding the experiences, preferences, and barriers to engagement of those affected by PD is essential for developing personalized, long-term interventions that enhance walking and reduce fall risk. Therefore, in this study we will further evaluate for whom and why daily life gait improves after the training intervention.

Even when assessed on the treadmill, despite a reliable average improvement in gait performance in PwPD, substantial inter-individual variability exists in training responses [44]. Apart from individual characteristics, the observed discrepancies in clinical outcomes may also, at least partly, be attributed to the heterogeneity of treadmill interventions. For example, to mimic the adaptive gait demands of daily life, some intervention studies added virtual reality and external perturbations to their protocols [45–47], which we refer to as “SDTT+ protocols”. Variations in SDTT+ type, the duration and the amount of training sessions may all affect training outcomes. For example, one study found that reactive postural control improved in ∼44 % of PwPD after treadmill training with mechanical perturbations versus ∼10 % in treadmill training without perturbations [44].

In the current multi-centric intervention study (StepuP) we will investigate treadmill training effectiveness and underlying biomechanical and neurophysiological mechanisms across SDTT and SDTT+ training protocols.

First, we will replicate that:

- Speed dependent treadmill training improves gait performance as indicated by an increase in gait speed following the training. (H1)

Second, regarding the biomechanical and neural mechanisms we hypothesize that:

- treadmill training improves gait performance through improved stability-related foot placement control. (H2)
- improved foot placement control correlates with changes in cortical activity and corticomuscular connectivity reflecting improved sensorimotor integration. (H3)

We further aim to understand:

- the added value and potential mechanistic effects of incorporating additional task demands into treadmill-based gait training.
- whether improvements in gait in supervised conditions translate to daily-life gait quantity and quality, and through which factors this is mediated.
- individual responses to treadmill training in the context of their neuromuscular control, clinical ratings, and individual characteristics.

## Methods

The “Steps against the burden of Parkinson’s Disease” (StepuP) consortium was funded by the JPND (Joint Programme – Neurodegenerative Disease Research) 2022 call for research on understanding the mechanisms of non-pharmacological interventions. The consortium consists of six partner sites: ETH Zürich (Zurich, Switzerland), IRCCS Istituto delle Scienze Neurologiche di Bologna (Bologna, Italy), Kiel University (Kiel, Germany), Tel Aviv Sourasky Medical Center (Tel Aviv, Israel), University of New South Wales (Sydney, Australia) and Vrije Universiteit (Amsterdam, Netherlands). It joins methodologies from biomechanics, neurophysiology and biomedical engineering with clinical expertise.

### Study design

The project is divided into four work packages: Randomized controlled multicenter treadmill training trial (RCT); Mechanisms underlying gait improvements after treadmill training; Daily-life mobility changes after treadmill training; and PwPD engagement and understanding individual responses to treadmill training.

The RCT is conducted in Bologna, Kiel, Sydney and Tel Aviv. A total of 126 PwPD will be enrolled across Bologna (n=21), Kiel (n=42), Sydney (n=42), and Tel Aviv (n=21), and blindly randomized into an intervention group (IG) and a control group (CG) by site. All CG participants will receive speed dependent treadmill training (SDTT) aimed at improving gait. Participants in the IG will receive the same SDTT with additional components (SDTT+). Baseline (T0) and follow-up (T1 & T2) visits will include descriptive, clinical, kinematic, and neurophysiological assessments and a 7-day home monitoring with Inertial Measurement Units (IMUs). In Sydney, half of the IG and CG participants will undertake two baseline measurements (T-1 & T0) before receiving the SDTT to evaluate measurement reliability.

At the Vrije Universiteit Amsterdam (Amsterdam, Netherlands) 21 age and gender matched healthy adults will undergo the T0 assessments as a reference group. The matching procedure will be performed with the participants recruited across the different clinical centers, ensuring representativeness of the respective cohorts.

#### Study timing

Each participant will undergo a total of five phases in the following order (Fig 1):

- *Pre-training*: The pre-training phase, which includes screening, enrolment, real-world assessment and functional assessment (T0) will last two weeks. A maximum of 14 days between the enrolment and the start of the first training visit is tolerated. In Sydney, half of the PwPD who are randomized into the control group and half of those who are randomized into the intervention group, will undertake the second baseline measurement (T-1, T0); thus, the pre-training phase in Sydney will be extended by 6 weeks ± 1 week.
- *Training*: The SDTT will consist of 12 sessions in total. Between training sessions a break of 0-4 days is planned, depending on the state of the participant and the logistics at each clinical centre.
- *Post-training*: The post-training phase, including real-world assessment and functional assessment (T1), will last for a total of 14 days. The functional assessment is planned no later than 2 days after the last training visit. The real-world assessment starts no later than 5 days after the functional assessment.
- *Maintenance period*: All participants will be offered an app-based real-world speed dependent walking training intervention.
- *Follow-up*: The follow-up visit (T2) will include a real-world assessment and a functional assessment and will take place 12 weeks (±2 weeks) after the completion of the post-training phase.

**Fig 1.**
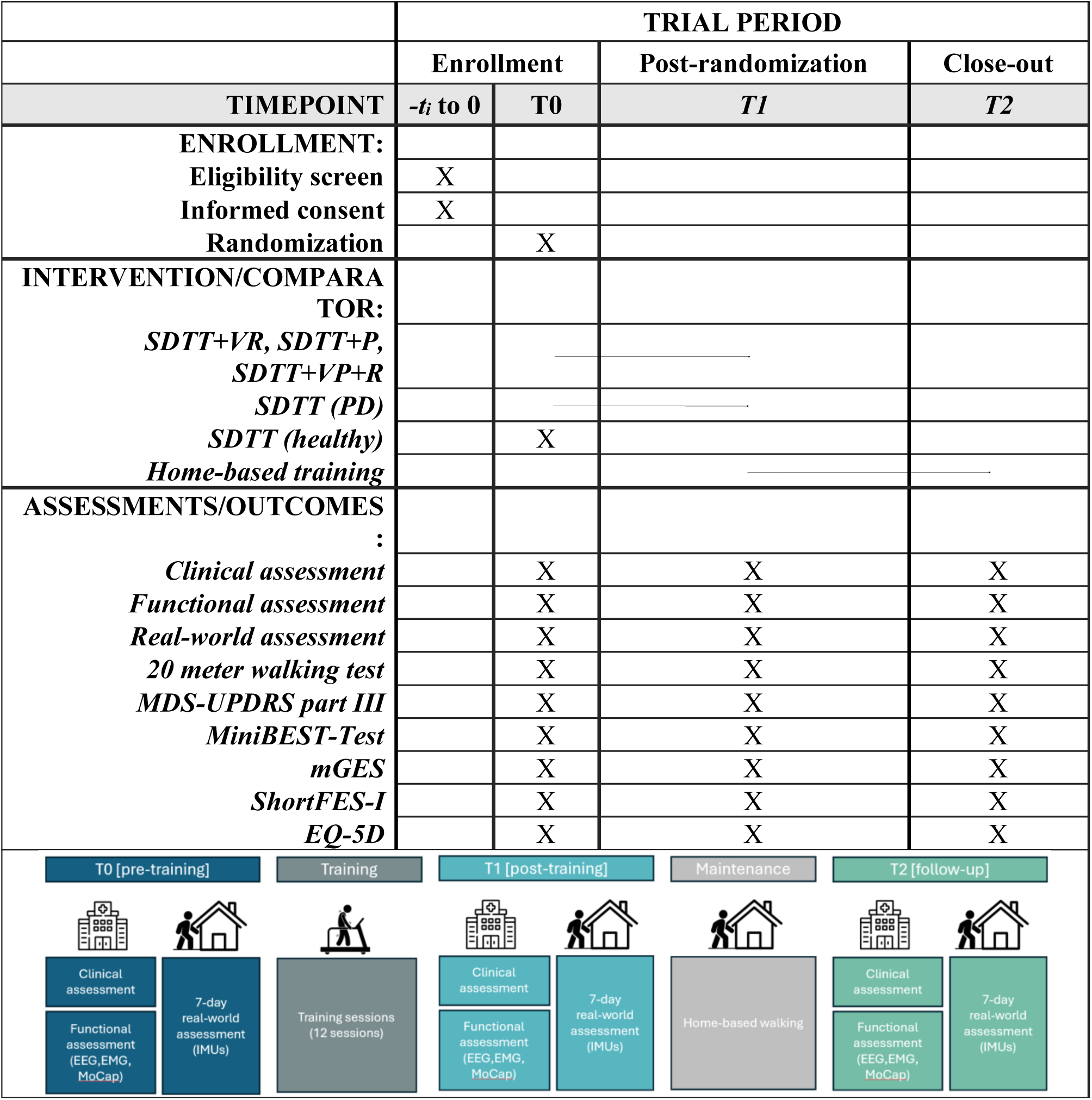
Participant timeline: schedule of enrolment, intervention, and assessments. EEG, elecroencephalogram; EMG, electromyography; FACIT-F, Functional Assessment of Chronic Illness Therapy–Fatigue; FES-I, Falls Efficacy Scale–International; IMU, Inertial Measurement Unit; MDS-UPDRS, Movement Disorder Society-sponsored Unified Parkinson’s disease Rating Scale; mGES, modified Gait Efficacy Scale; MiniBEST, Mini-Balance Evaluation Systems Test; MoCap, motion capture.

Recruitment has started on the 1st of August 2024 and is expected to be completed by the 31st of March 2026. Data collection has commenced in August 2024 and is expected to be completed in October 2026. Results are expected from March 2027.

#### Ethical approval

Ethical approval was obtained at the individual sites (Kiel D 629/24, Tel Aviv 0569-23-TLV, Sydney iRECS5114, Bologna 71-2024-SPER-AUSLBO, Amsterdam VCWE-2025-083). All participants will provide written informed consent and the research will be performed in accordance with the Declaration of Helsinki. The trial has been registered at clinicaltrials.gov (ids: NCT07057219, NCT07105787, NCT07058285, NCT06538909).

### Participants

Participants will be enrolled in the study across the four clinical centers in accordance with the inclusion and exclusion criteria detailed in Table 1. Recruitment will be conducted via registries and hospital records or via flyers and local advertisements.

For the healthy reference group in Amsterdam, a specific exclusion criteria questionnaire (S1) will be applied, ensuring that all participants in this group do not have any (neurological) impairments or injuries which could affect their walking pattern.

**Table 1.**
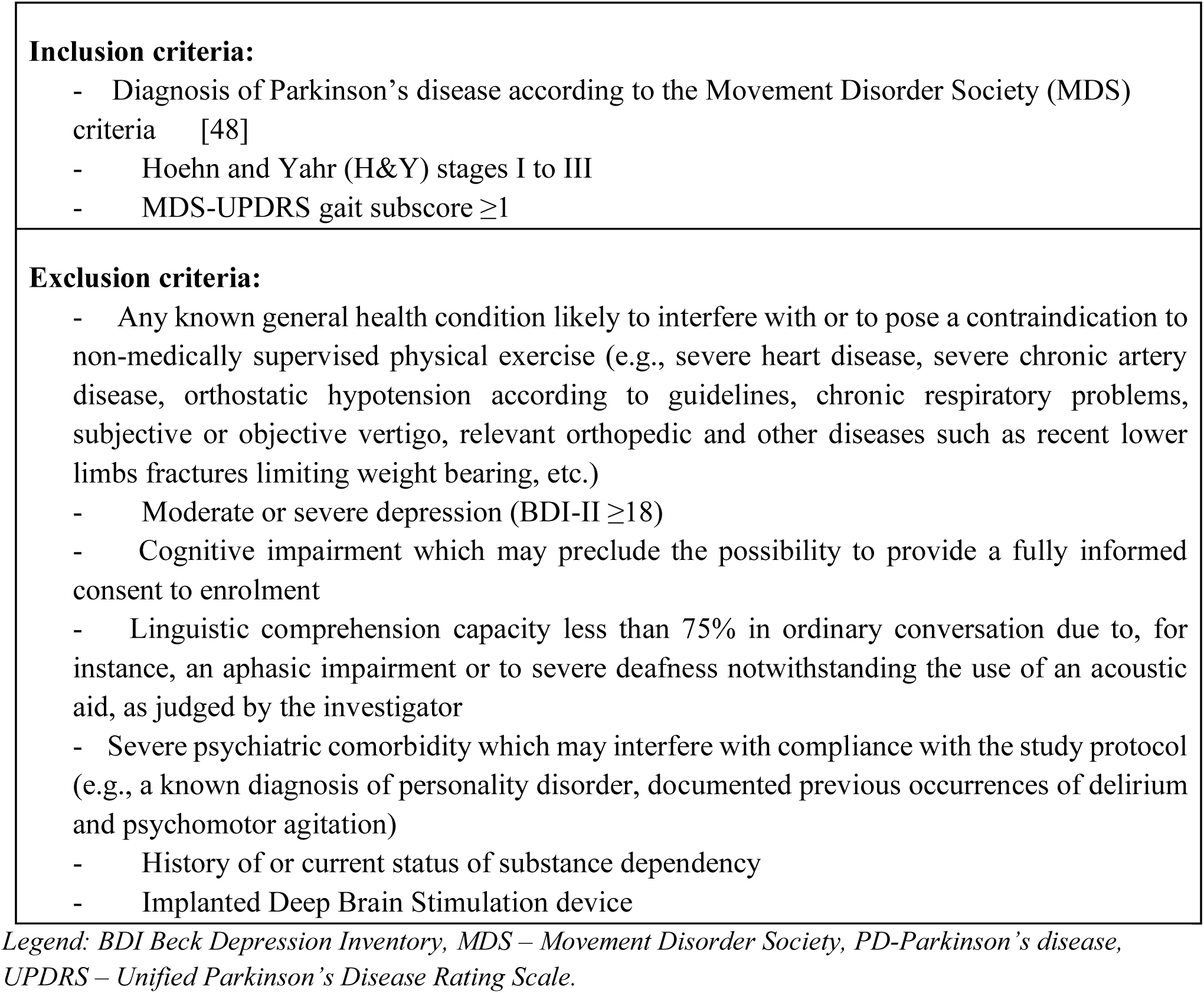
Inclusion and exclusion criteria for participation in the StepuP trial.

#### Randomization procedure

A researcher not located at a site where recordings of PwPD are performed (M.v.L. in Amsterdam) will prepare randomization sequences using a custom Python script with block randomization stratified by site and H&Y stage and securely uploaded to REDCap. The block sizes will randomly range between 2 and 4 for each of Bologna and Tel Aviv, and 2 and 6 for each of Kiel and Sydney. To ensure allocation concealment, participants will be randomized into the SDTT or SDTT+ group after the T0 assessment. This ensures that neither recruiters nor participants will be aware of group assignments beforehand, thereby minimizing the risk of selection bias.

### Intervention and control

All participants will undertake a treadmill-based intervention, comprising 12 sessions of 30 minutes each.

To understand the effects of additional components, each center will conduct SDTT and SDTT+. Tel Aviv and Bologna will apply virtual reality (VR), while Kiel will administer mechanical perturbations, and Sydney will apply both VR and mechanical perturbations (Table 2). The SDTT+VR program performed in Bologna and Tel Aviv will use the Gait Better system [21], where the participant is required to step over obstacles which appear on the screen eliciting proactive gait adaptations. The SDTT+ perturbations program performed in Kiel will employ accelerations and decelerations of the treadmill belt to elicit reactive gait adaptations. The SDTT+ program in Sydney involves both VR and perturbations where the participant is required to step over virtual obstacles and treadmill belt accelerations or decelerations occur when obstacle-foot collision occur [49], eliciting both proactive and reactive gait adaptations.

**Table 2.**
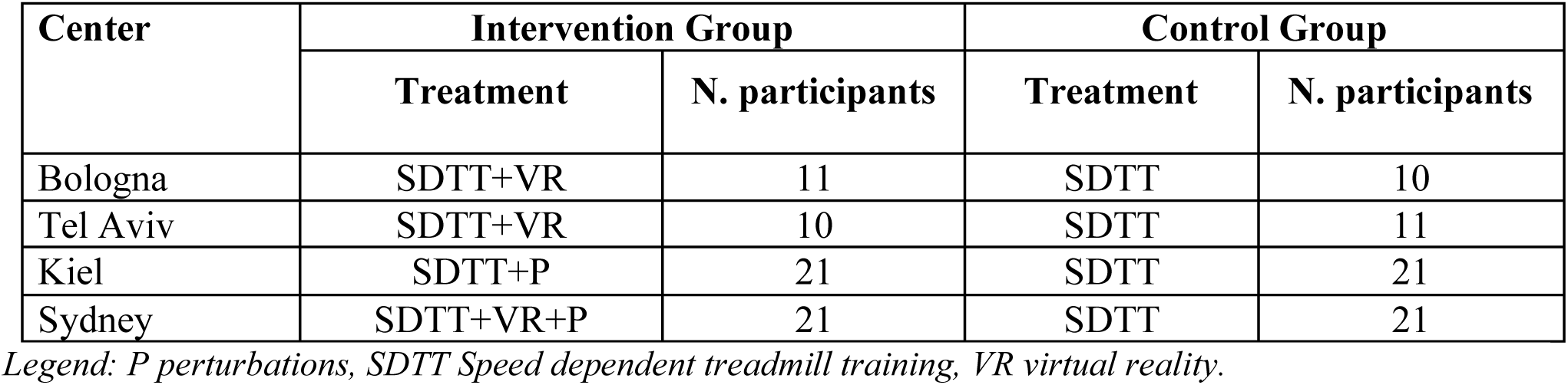
Intervention and Control groups.

#### Treadmill training protocol

At the first training of each training week, a single 20-meter walk test will be performed before the treadmill training in order to define the comfortable walking speed for training (CST). The CST will be used to individually tailor treadmill training intensity, ensuring that the intervention is adapted weekly to the current functional capacity of each participant.

Each training session consists of three training blocks at three different speeds (V1, V2 and V3), intermingled with recovery trials. V1 will always be set at 80% of CST, whereas V2 and V3 may progress from 80% to 110% of CST according to the training progression protocol (Table 3). Each session begins with a warm-up period (walking at 70% of CST). Between training blocks is a recovery phase where participants walk at 70% of CST for 2.5 minutes.

During training, participants will wear an overhead safety harness to prevent falls. Treadmill railings may be held only if needed and only during the first minute of each training block to allow the participant to adjust to the novel speed demands. Then, participants will be encouraged to use reciprocal arm swing.

All training sessions will be conducted individually to ensure safety and to allow close supervision of each participant. A physiotherapist will be present to monitor the participant’s gait pattern and signs of fatigue but will not give any physical assistance during the walking. Furthermore, the physiotherapist will ensure that adequate rest breaks are inserted to maintain participant safety and will record their frequency and duration.

**Table 3.**
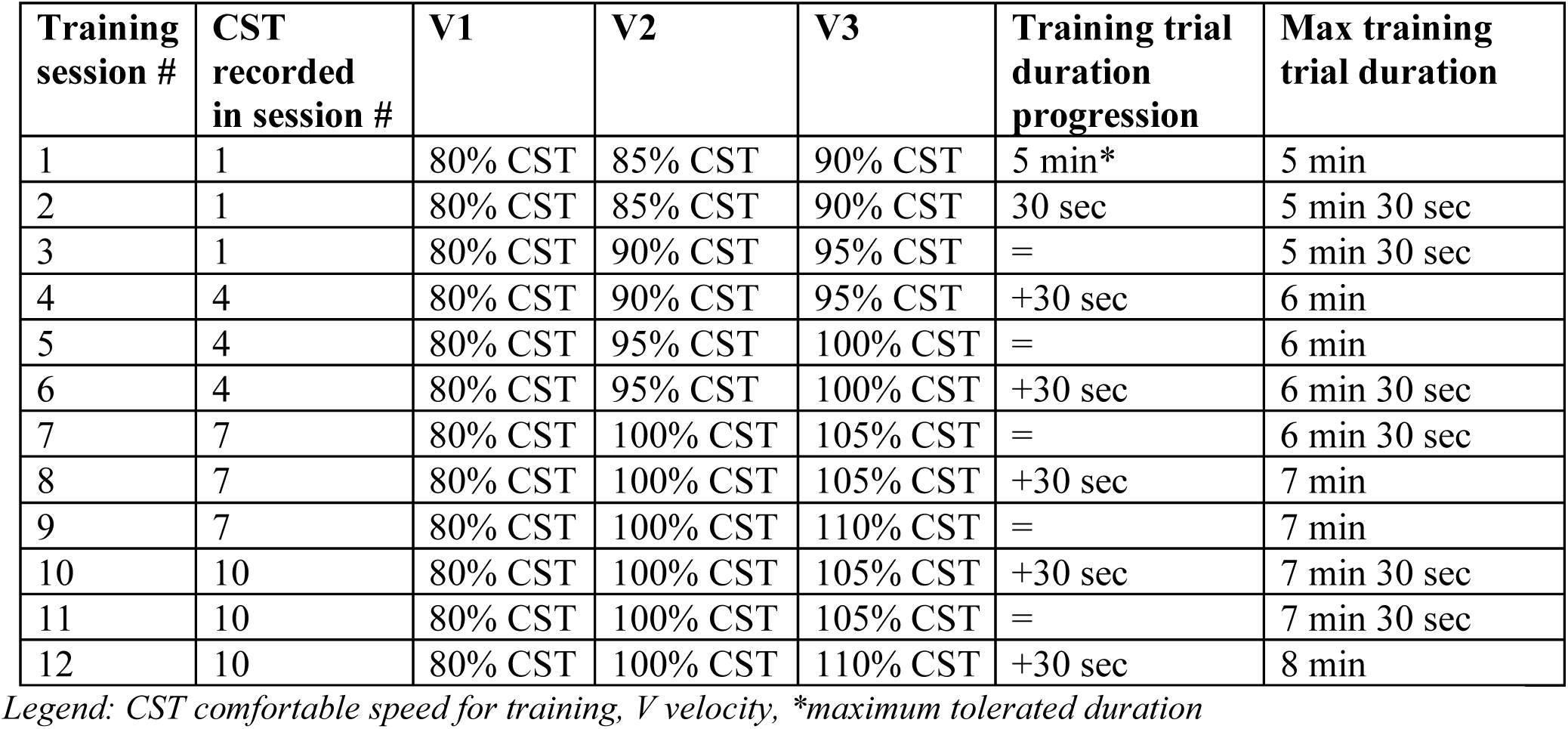
Progression of training speed and duration.

#### Training progression

A physiotherapist or exercise physiologist will adapt the training progression according to the participants’ level of exertion and performance. Speeds of V2 and V3 will be only increased by 5% if the previous maximum attained speed was tolerated. If the speed was not tolerated, it will be reduced by 5% to the previously tolerated walking speed before the bout is resumed. If the speed was tolerated but not the duration of the walking block, the duration of V2 and V3 will be shortened (- 30 seconds) to the previously tolerated walking block duration. Additionally, a longer (+30 seconds) recovery phase (walking at 70% CST) or a seated rest may be included if required.

In addition, perturbation intensities may be increased. Treadmill belt accelerations and decelerations which induce forward and backward balance loss, respectively, start at ±4 m/s^2^ for 100 ms which may be increased to ±15 m/s^2^ for 200 ms. The VR challenge starts with obstacles with 10 cm height or 16 cm length and up to 20 cm height and 30 cm length, requiring higher and longer steps, respectively.

Personalization of the training progression including speed and during of training blocks, recovery phases, total duration, and VR/P intensities will be recorded.

#### Home-based intervention

During the maintenance period (from T1 to T2 assessments), all participants will be offered a home-based speed dependent walking training intervention using the Walking Tall app (https://walkingtallhealth.com.au/). The mobile app (available for both Android and iOS devices) delivers a rhythmic metronomic beat for three different walking speeds, designed to trigger movement and encourage better walking patterns. Participants will be supported using the app to progress towards World Health Organisation recommendations for moderate-intensity exercise through a personalized goal-setting session.

### Outcome measures

#### Clinical assessment

At each assessment (T0,T1,T2), a standardized clinical evaluation will be conducted using a Case Report Form (CRF) to characterize the participant on a clinical level. Initial assessments include demographics, medical history and medication use. Cognitive function, motor performance, and mobility will be assessed through a combination of clinical and functional scales, as well as instrumented tests, to ensure a comprehensive evaluation. Specifically, cognitive function will be assessed using the Montreal Cognitive Assessment (MoCA) [50] and the Color Trail Test (CTT) [51]. Motor performance will be measured with the MDS-Unified Parkinson’s Disease Rating Scale part 3 (MDS-UPDRS-III) [52], postural control and fall risk will be assessed with the Mini-Balance Evaluation Systems Test (MBT)[53], walking capacity with the 2-minute walk test (2MWT), mobility performance with the Timed Up and Go (TUG) test, and confidence in walking abilities with the modified Gait Efficacy Scale (mGES) [54,55].

Fear of falling will be assessed with the Falls Efficacy Scale–International (FES-I) [56], fatigue with the Functional Assessment of Chronic Illness Therapy–Fatigue (FACIT-F) [57], health-related quality of life with the EQ-5D-5L [58], pain perception with the Visual Analog Scale (VAS) [59], freezing episodes with the New Freezing of Gait Questionnaire (NFOGQ) [60], and depressive symptoms with the Beck Depression Inventory–II (BDI-II).

At T1 and T2 assessments, the assessors will be blinded to the type of training performed by the participants.

#### Functional assessment

Comfortable walking speed will be assessed through the 20-meter walk test (which will then be used as a starting point for treadmill-based assessments).

Motion, electromyography (EMG), and electroencephalography (EEG) data will be recorded during treadmill walking. The motion capture data will allow quantification of the quality of foot placement control. The quantification will be based on a linear foot placement model [25], describing the relationship between the center-of-mass kinematic state during swing and foot placement at the end of the swing phase. The EEG and EMG data will be combined to assess corticomuscular coherence reflecting sensorimotor integration hypothetically related to this stability mechanism. Fig 2 shows the timeline of the functional assessment, which is described in detail in the following sections. The functional assessment takes about 1.5 hours to complete, including the participant preparation time (i.e. attaching the sensors etc.). Prior to any motor assessment the sensors used for the daily life recordings will be attached.

**Fig 2.**
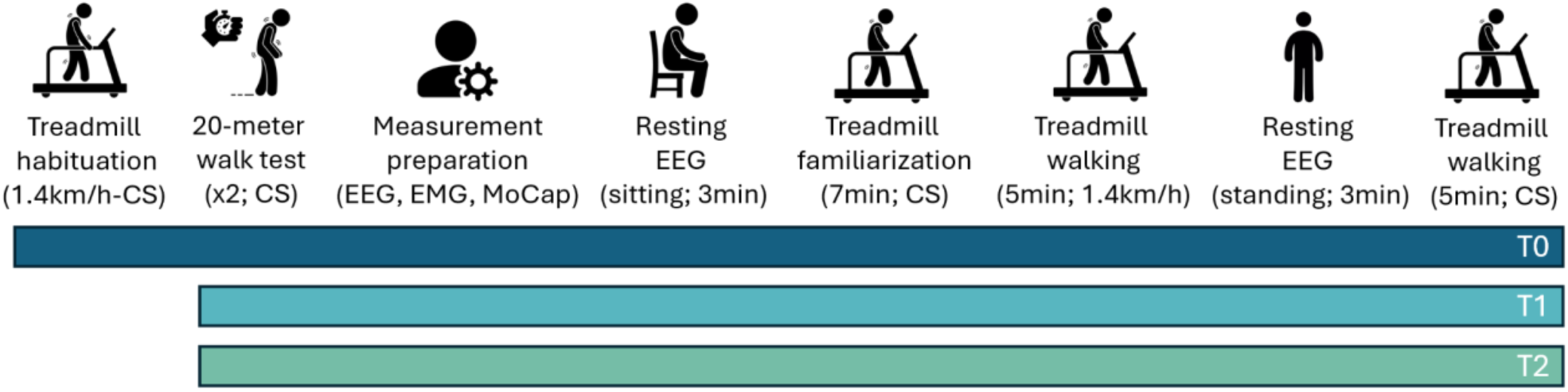
Functional assessment timeline. The tests performed at T0,T1, and T2 are indicated by the blue, light blue, and green lines, respectively, shown at the bottom of the figure. For each recorded task, additional information is reported in parentheses (e.g., duration, standing/sitting condition for static tasks, and velocity for dynamic tasks).

##### Treadmill habituation

At T0, the participant will have up to 7 minutes to get used to treadmill walking. The habituation will be completed once the participant feels comfortable walking on the treadmill, whilst wearing a harness, but without handrail support. If during the habituation the participant is unable to walk at 1.4 km/h but is able to walk at their preferred walking speed on the treadmill, the participant will still be eligible to participate in the study. In this case, their preferred walking speed at T0 will be repeated at T1 and at T2 instead of the 1.4 km/h, to ensure one standard speed remains.

##### 20-meter walk test

Comfortable walking speed (CS) will be assessed using a 20-meter walk test. Participants will perform four trials of the 20-meter walk test (two as “familiarization” to the task, and the last two to be averaged over as an outcome measure) [61]. Between the first two and last two trials there will be break of approximately five minutes depending on the state of the participant. A 24-meter walkway with a start and finish line, clearly distinguishable by tape on the floor will be used. A 2-meter mark will be placed respectively 2 meters from the start and finish line, to aid the experimenter in accurately timing the 20-meter walk. The 24-meter walkway will not contain any door entrances or other environmental factors, such as narrow passages, that could trigger freezing of gait. The assessor will start the stop watch when the participant crosses the first 2-meter mark and stop the timer once the first heel of the participant has crossed the 2-meter mark before the finish line.

The participant will be instructed as follows:

“Now you will walk from this line to that line. Try to imagine walking a longer distance at a speed that is comfortable for you. There is no need to rush, it is not a competition, you are not running for the bus. Please start behind this line and make sure you continue walking across the finish line at the other side until we say STOP.”

##### Measurement preparation

After the 20-meter walk test, motion capture markers, the EEG cap and EMG electrodes will be placed. In addition, a video of the participant’s head will be recorded whilst wearing the EEG cap and sitting still (see “Data collection” below for more detail).

##### Resting state EEG measurements

Two eyes-open resting state EEG measurements will be performed: one before treadmill walking, and another one after familiarization. During the first resting state measurements, participants will sit in a chair, arms resting on their lap, and will be instructed to remain as still, and silent, as possible for three minutes, whilst fixating their gaze on a clearly visible fixation cross on the wall. The second EEG resting measurement will be performed while standing with the arms relaxed on the sides of the body for three minutes. The resting state EEG measurements will be used as a reference, containing no gait-related movement artefacts.

##### Treadmill walking

During all treadmill trials participants will wear an overhead harness, which does not support the participant’s weight, but prevents the participant from falling. Someone with familiarity and expertise with the assessments (e.g. a physiotherapist) will be present to monitor the participant but will not provide physical assistance.

Participants will walk on the treadmill at CS for seven minutes as familiarization (familiarization time required for gait parameters to stabilize [62]), followed by a five-minute break, during which resting state EEG will be recorded. After which they will walk one five-minute trial at 1.4 km/h and one five-minute trial at their CS, with a 5-minute break in between.

In case of time constraints, the 1.4 km/h trial will always be prioritized over the CS trial. For participants who could not achieve 1.4 km/h at T0, this is the trial with their individualized standard speed, to be repeated at T0, T1 and T2.

#### Real-world assessment

Following each functional assessment (T0, T1, and T2), unsupervised data will be collected during daily life over seven consecutive days using IMUs with a sampling frequency of 100 Hz (gyroscope range: ±2000°/s, accelerometer range: ±8 g) and reduced dimensions (23×32.5×8.9 mm; mass 11 g). Participants will wear an IMU (Axivity AX6, Axivity, Newcastle upon Tyne, UK) on the lower back at the fifth lumbar vertebra. During the functional assessment, the IMU will be secured with two layers of adhesive medical tape.

In a volunteering subgroup of participants, two additional sensors will be attached to the shanks, positioned above the lateral malleolus, to investigate the coordination between the CoM and the feet during daily life and evaluate the differences in spatio-temporal gait outcomes collected from different body positions (lower back and shanks). These sensors will be secured using a neoprene ankle band, which participants can remove (e.g., during shower), reattach, or adjust for width.

During the real-world assessment period, the devices will be worn continuously up to 7 days, including at night, as they do not require charging (i.e., the battery lasts more than 7 days). Participants can maintain their usual activities, such as sports or showering, since the chosen sensors are waterproof. The sensor can be removed if wearing them becomes uncomfortable or if it causes irritation to the skin. The participants will be provided with detailed information on how to reattach the sensor and additional adhesive medical tape. Data will be stored locally on the IMU. After the seven-day monitoring period, participants will return the sensors using a provided return envelope, and the data will be downloaded.

##### Diaries

During the 7-day monitoring periods at T0, T1 and T2, participants will complete daily paper-based diaries to record medication intake, physical activities, freezing of gait (FOG) episodes (number of FOG occurrences “before/after lunch”), and falls. The diaries will cover the period from 6 AM to 10 PM each day from day 1 to day 7.

In the medication diary, participants can either mark their scheduled medication intake or note any deviations for that day. In the physical activity diary, participants will indicate when they carry out specified activities each day (e.g., walking, swimming, physiotherapy). In addition, to assess activity levels during the real-world assessment, participants will complete a questionnaire at the end of the 7-day period (S2). This questionnaire will include questions about the use of walking aids, the perceived “normality” of the week, and levels of mental and physical fatigue during the real-world assessment. The completed diaries will be returned along with the IMU at the end of the monitoring period.

#### Understanding Engagement, Individual Response, and Future Improvement

To understand why treadmill training benefits some individuals with Parkinson’s disease (PD) more than others, we will integrate qualitative and quantitative data across sites. Qualitative data will be collected from all participants through online questionnaires, including the System Usability Scale (SUS)[63], Physical Activity Enjoyment Scale (PACES) [64], and Exercise Self-Efficacy Scale (ESES) [65], each accompanied by free-text fields for feedback on barriers and enablers to long-term use. In addition, a stratified subsample of participants from each intervention arm and site will complete semi-structured exit interviews at T2 [66].

### Data collection procedures

#### Kinematic data

Kinematic data will be collected using a Qualisys (in Amsterdam, Kiel) or Vicon (in Bologna, Sydney, and Tel Aviv) motion capture system at a sample frequency of 100 Hz.

Heel, pelvis and one additional asymmetry marker on the right foot comprise the most minimal marker set needed for the main analysis (Table 4). At some sites, the Vicon plug-in gait model is used to extend the marker set, and gain more information about the gait pattern.

**Table 4.**
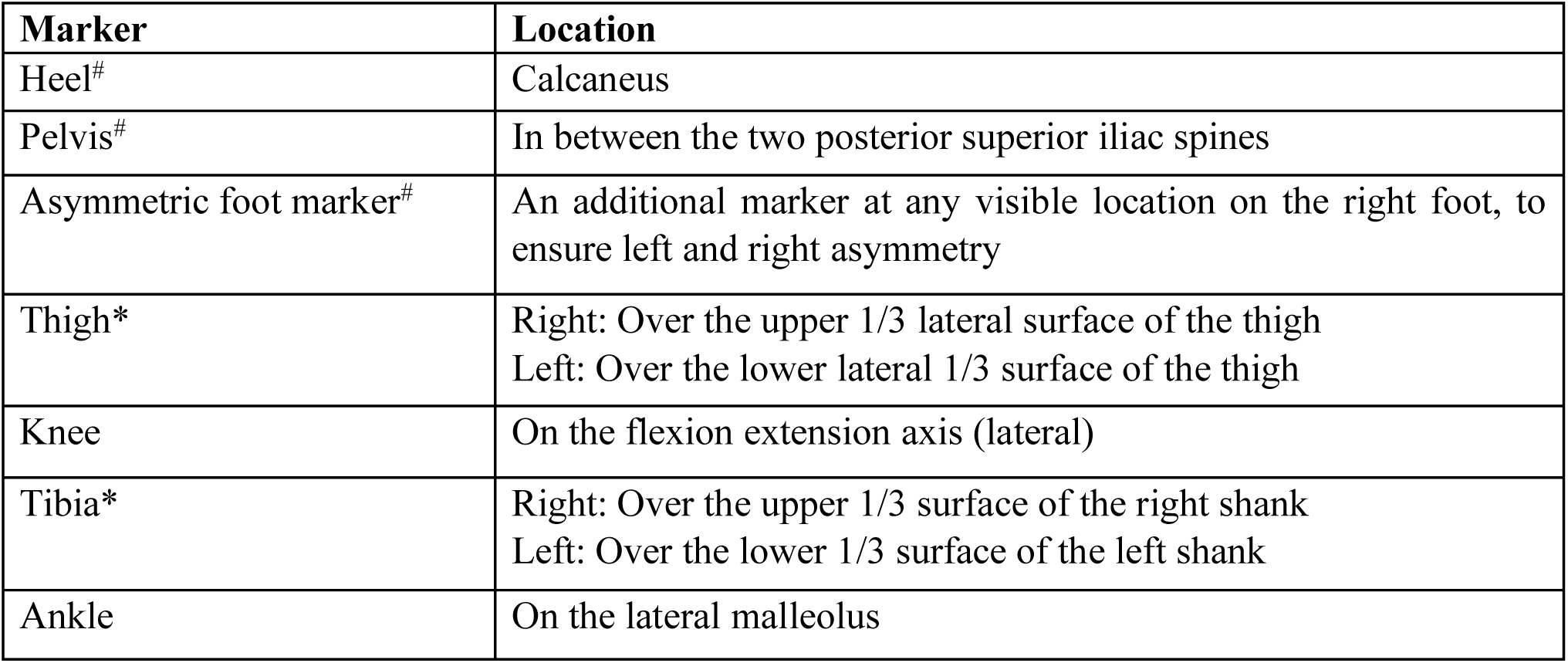
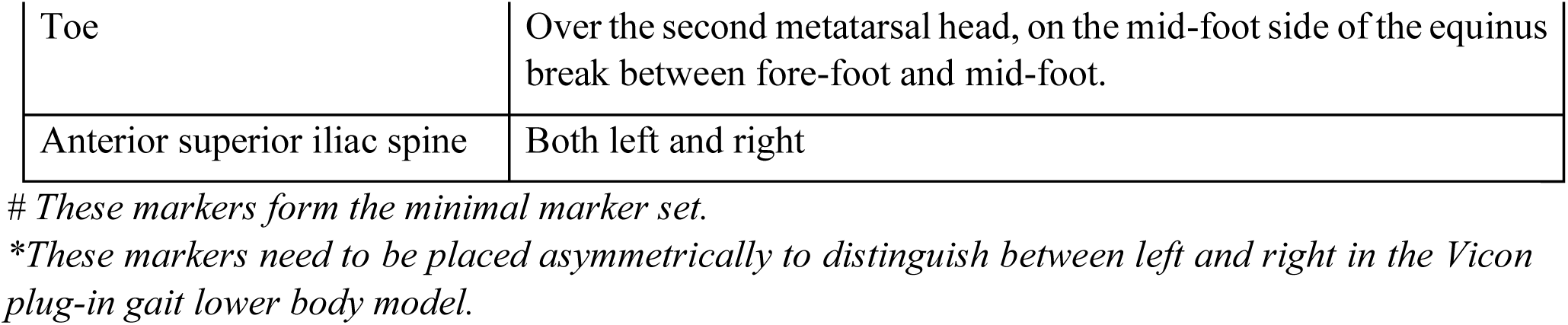
Placement of motion capture markers.

#### Electromyography

Electromyography data will be recorded using Cometa Pico EMG (in Bologna) or Delsys Trigno EMG (in Amsterdam, Kiel, Sydney, and Tel Aviv) at a sample frequency available for each system around 2148 Hz.

Bipolar electrodes will be placed on the leg muscles as listed in Table 5, the first three muscles listed will be measured at each site, whereas the other muscles are optional in this protocol. EMG electrodes will be placed in accordance with the Seniam guidelines for electrode locations (http://www.seniam.org/). Hair will be shaved, and the skin will be cleaned with alcohol prior to attachment of the electrodes. After applying all the electrodes, the EMG signal will be checked by asking the participant to contract the muscles briefly (e.g., abducting the leg a couple of times for gluteus medius muscle, and rotating the ankle a couple of times for the ankle muscles). The power spectrum will be checked for line noise, and precautions will be taken to limit this noise.

**Table 5.**
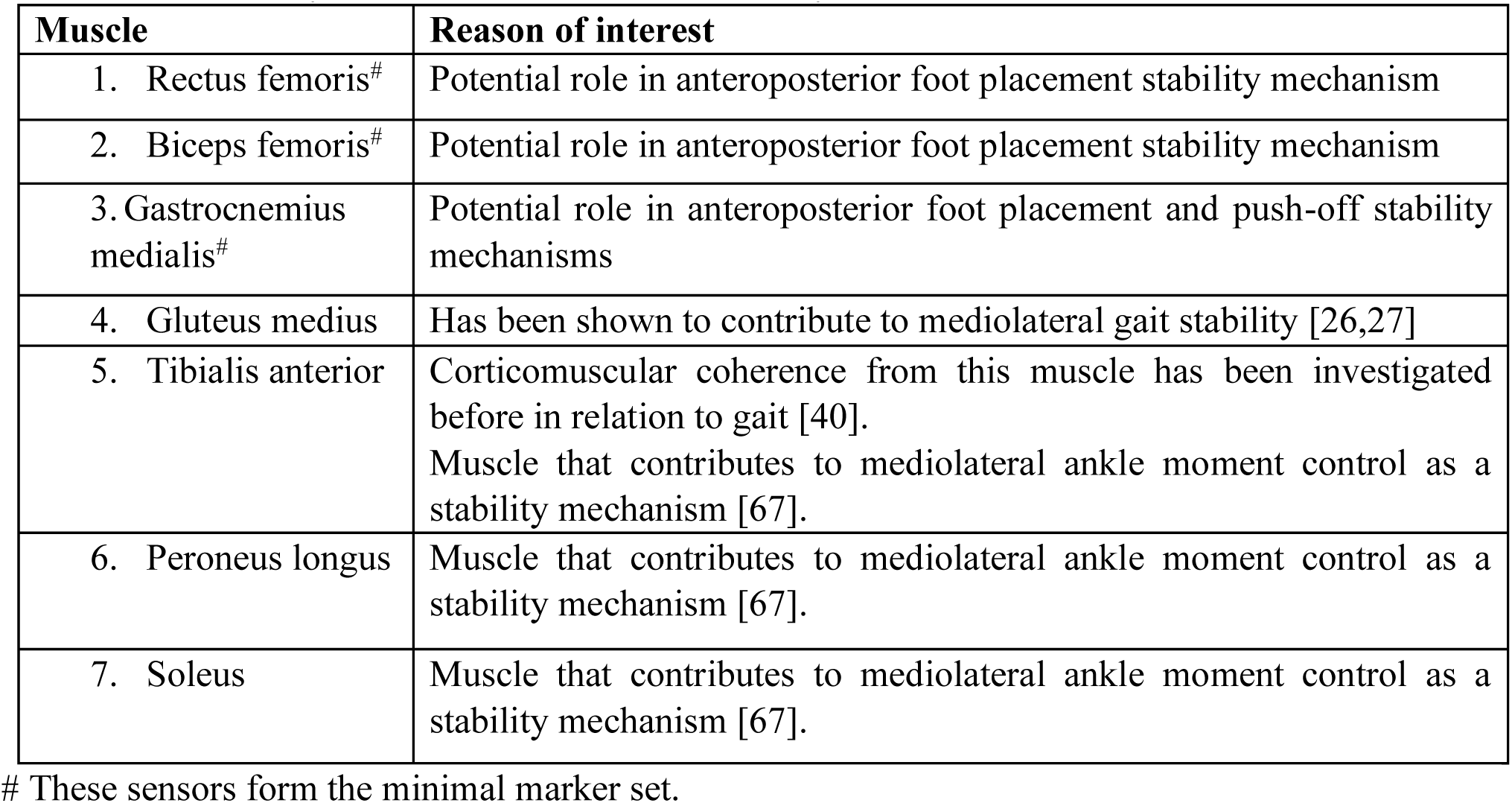
Placement of EMG sensors and their reason of interest.

#### Electroencephalography

Participants will be asked to come to the lab without hair gel or other products in their hair, to avoid effects on electrode impedance. Depending on the sites preferences and facilities the hair will be washed and dried upon arrival to further mitigate or reduce impedance. Jewelry such as earrings or piercings will be removed prior to the recording.

EEG data from PwPD will be collected using a Smarting X Pro device (in Bologna, Kiel, and Sydney) or a g.Nautilus wearable EEG headset (in Tel Aviv). Both devices are fully wearable and wireless to maximize participant comfort during treadmill walking, ensuring freedom of movement and data recording without wire interference. In Amsterdam the TMSi Saga device will be used for EEG data collection from healthy control participants. This system will be used wired in a way that minimizes interference with the walking pattern and comfort of the participant.

All recordings will be performed using a 64-channel EEG cap. The size of the cap will be chosen based on the measured circumference of the participant’s head. Caps will be placed in accordance with the 10-20 layout. To this end, we will ensure the Cz channel is positioned at the crossing of lines between nasion and inion and the two preauricular points. The exact positioning of the cap on the participant’s head will be registered through a video recording made with a mobile device. This information will help us to source-localize brain activity in our analyses. For such purposes the fiducials are used as references to the head. Therefore, the fiducials will be marked e.g. with small stickers clearly distinguishable in the video. To capture video footage for Neural Radiance Fields reconstruction of the participant’s head, we will ensure the camera is set up in a well-lit environment with a plain background, and maintain focus on the participant’s head throughout the recording. The video will be recorded around the participant’s head, whilst making sure all sides are covered.

Electrode-skin contact will be established by using cotton swabs and abrasive/conductive gel. Target impedances are below 10 kOhm. Impedances will be monitored during cap preparation and digitally registered before and after EEG measurements.

Two fixation crosses will be placed on the wall, corresponding to eye level in upright and sitting positions, respectively. Participants will be instructed to focus on the fixation cross during walking and resting measurements.

#### Synchronization procedure of multimodal recordings

The synchronization at the clinical centers of multimodal physiological (EEG and EMG) and kinematic (optical motion capture) data streams will be achieved through the open-source Lab Streaming Layer (LSL) framework [68]. All synchronization scripts are publicly available (https://github.com/JuliusWelzel/StepuP_setup/tree/main/sync_scripts).

In Amsterdam a TTL pulse will be sent from the motion capture system (Qualysis), which will be recorded alongside the EEG signals and which will start and stop the EMG recording.

### Data management procedures

#### Data standardization (BIDS)

To ensure high standards of data processing, all data will be organized in line with the Brain Imaging Data Structure (BIDS) [69–71] and structured as follows: Raw time-series data from EEG, EMG, and motion capture are stored in modality-specific subdirectories (eeg, emg, motion) under participant (sub-) and session (ses-) folders, with filenames encoding task identifiers (e.g., sub-01_ses-01_task-treadmillComfortableSpeed_eeg.vhdr). Additional demographic and clinical data will be stored in the accompanying “participants.tsv” file.

#### Data access and storage

We will integrate qualitative and quantitative data across sites within a secure, General Data Protection Regulation (GDPR) compliant analysis environment. Participant data will be collected in a coded, de-identified manner. Data will be stored on a REDCap server in a standardized form across the StepuP consortium. All recruitment sites will keep original records of all signed consent forms, trial key codes, and any other paper forms or samples that are collected at source, under secure conditions.

### Endpoints

#### Primary endpoint

The primary endpoint will be comfortable gait speed during overground walking as assessed with the 20-meter walk test.

#### Secondary endpoints

##### Clinical endpoints

Secondary clinical endpoints are the EQ-5D, Mini-BESTest, mGES, Short FES-i, and MDS-UPDRS Part III. They will assess health status, functional abilities, and the impact of treadmill training on quality of life, balance, walking confidence, fear of falling, and motor symptom severity in Parkinson’s disease

##### Foot placement control

To quantify stability-related foot placement control, we will use the foot placement model as previously reported by Wang & Srinivasan (2014), (equation 1).

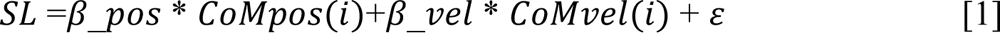

Prior to fitting this model, all relevant variables will be time-normalized from 0-100% of the swing phase (51 samples). SL is the demeaned actual step length of a particular step (measured as the anteroposterior distance between the two heel markers) and 𝜀 is the residual (i.e. the difference between the actual and predicted step length). 𝐶𝑜𝑀𝑝𝑜𝑠 and 𝐶𝑜𝑀𝑣𝑒𝑙 represent respectively the demeaned position and velocity of the center-of-mass (i.e. estimated as the pelvis marker) with respect to the stance leg for a percentage (*i*) of the swing phase, and 𝛽_𝑝𝑜𝑠 and 𝛽_𝑣𝑒𝑙 as their respective regression coefficients/gains. The model fitted for the center-of-mass kinematic state (i.e. 𝐶𝑜𝑀𝑝𝑜𝑠 and 𝐶𝑜𝑀𝑣𝑒𝑙) at percentages early on in the swing phase can be considered as representative of feedback control, whilst the model fitted for the center-of-mass kinematic state at the end of the swing phase as a performance measure of stability related foot placement control. Therefore, foot placement endpoints will be tested both for the model fitted for the midswing *(i = 25*) CoM kinematic state, as well as for the model fitted for terminal swing CoM kinematic state (*i = 51*).

The R^2^ measure of the foot placement model summarizes the degree of foot placement control (i.e. how well foot placement is coordinated according to the CoM kinematic state). The gains of the model (𝛽_𝑝𝑜𝑠 and 𝛽_𝑣𝑒𝑙) give insight into the strength of the foot placement responses. The standard deviation of the residual represents the magnitude of the foot placement errors and can be considered as a measure of foot placement precision.

Although the training interventions are focused on improving anteroposterior foot placement control, the model will also be fitted for mediolateral foot placement control (i.e. step width control) for exploratory purposes.

##### Beta power and Corticomuscular Coherence

To evaluate cortical involvement, and possible sensorimotor integration, underlying foot placement control, we will compute EEG spectral beta power and corticomuscular coherence in the beta frequency band (13-30 Hz). These measures will be calculated from source-localized EEG signals and EMG signals from muscles involved in foot placement control, primarily the rectus femoris muscle. This is the anteroposterior equivalent of the mediolateral gluteus medius muscle, which has previously been associated with stability related step width control [26,27].

EEG analyses will be conducted with the use of the Fieldtrip Toolbox [72]. EEG pre-processing will involve band-pass filtering (0.5-220 Hz) and 50-Hz notch filtering to reduce line noise followed by Independent Component Analysis (ICA) based artefact removal. The principal artefacts expected are due to movement, head and neck muscle activity, ocular activity (eye movement, blinks), and ECG and will be extracted based on (adjusted) criteria from previous studies [32,73]. Following the removal of artefact components, an additional high-pass filter at 5 Hz will be applied. The data will be epoched in strides defined from heelstrike to heelstrike. Corticomuscular coherence will be computed across the gait cycle by means of time-frequency analyses followed by time-normalization of EEG and EMG cross-spectra. Time-normalization of each epoch will be done between gait events (i.e. from ipsilateral heelstrike to contralateral toe-off, from contralateral toe-off to contralateral heelstrike and from ipsilateral toe-off to ipsilateral heelstrike), as such that each stride consists of 101 time points, representing 0-100% of the gait cycle. The specific percentages between each gait event will be determined as the average percental instance of the gait event averaged across all participants and conditions [32]. Dynamic Imaging of Coherent Sources (DICS) beamforming will be used for source-localization of coherence in the beta band (13-30 Hz), time-averaged across the gait cycle. For accurate forward modelling we will use the electrode positions extracted from the video recordings. Source-localization will be performed separately for the corticomuscular coherence of the right and left muscles of interest. We will reconstruct the EEG time-series for the location of the identified sources and compute the time-dependent beta power and coherence that will be used as outcome measures in the statistical analyses. For foot placement control the time window of interest of these outcome measures will be around early-swing, whereas for ankle moment control the outcome measures will be considered during the stance phase.

##### Real-world assessment

To assess the extent to which gait improvements translate to daily life, we will evaluate both gait quantity and quality [74,75]. The primary endpoints of interest include steps per day, the amount and length of uninterrupted walking durations, gait speed, stride time variability and gait symmetry. These measures will help us identify for whom daily-life mobility improves following the training and for whom it does not. Additionally, the endpoints from the self-reported diaries will provide further insights into this understanding.

##### Assessment of Engagement, Individual Response, and Future Improvement

To identify characteristics of responders versus non-responders, machine-learning models will combine demographic, biomechanical, neurophysiological, and digital mobility data with coded qualitative features. Qualitative themes will be transformed into categorical or binary variables and, where appropriate, embedded using natural-language processing approaches.

To identify determinants of successful treadmill training and inform personalized, long-term strategies for improving mobility and preventing falls in people with PD, semi-structured interviews [66] will be conducted to explore experiences, preferences, and barriers/facilitators to treadmill training and use of the Walking Tall app. Transcripts and free-text responses will undergo reflexive thematic analysis to identify key themes related to acceptability, usability, motivation, and contextual factors influencing engagement and outcomes. Coding will be conducted iteratively by multiple researchers, with a proportion of transcripts double-coded to enhance analytic rigor and allow development of a shared codebook. Themes emerging from interviews and questionnaires will be synthesised into a prioritised list of modifiable barriers and enablers, mapped to specific intervention components. A summary of participant-driven recommendations and their implications for personalized gait rehabilitation will be reported to support future implementation and scale-up.

### Statistics

#### Sample size estimation

The sample size was calculated for the clinical primary objective (i.e., comfortable gait speed overground, measured in m/sec from a 20-meters walking test [76]), with a significance level (alpha) of 0.05 and power (1-beta) of 0.80, with an allocation ratio of 1:1 in parallel groups. From the available literature, it is known that SDTT may improve comfortable overground gait speed up to 0.1 m/sec (from a baseline of 1.26 m/sec) [77] in comparison to standard overground walking and that SDTT with perturbations (SDTT+) may further increase comfortable overground gait speed up to +0.08 m/sec in comparison to SDTT. Thus, we will assume that SDDT+ is superior to SDTT at improving comfortable overground gait speed based on an expected average difference in gait speed between the two groups of +0.08 m/sec with a SD of 0.08 [77,78]. Under this assumption, 17 participants per group are expected to be enrolled, rising to 21 considering a maximum drop-out rate of 20%. This would lead to 42 subjects per trial. Consequently, 126 participants will be enrolled in total.

#### Analysis plan

Statistical inference in the StepuP trial is anchored in pre-specified mixed-effects models evaluating changes in clinical, biomechanical, and neural outcomes over time. Mediation analyses are restricted to a small number of theoretically motivated pathways (e.g., gait efficacy mediating the relationship between lab-based improvements and daily-life mobility) and will be interpreted cautiously given sample size and site stratification.

Machine-learning analyses are explicitly exploratory and hypothesis-generating. Their purpose is not to maximize predictive accuracy, but to identify multivariate patterns and candidate moderators of training response across biomechanical, neurophysiological, clinical, and experiential domains. To mitigate overfitting, models will be constrained, evaluated using nested cross-validation, and interpreted primarily through transparent feature-importance metrics rather than classification performance. Findings from these analyses will be used to inform future confirmatory studies rather than to draw definitive causal conclusions. Collectively, these analyses prioritize mechanistic insight and internal coherence over exhaustive model complexity.

#### Main analyses

The main analyses are outlined below. For a more detailed overview of the endpoints and participants included for each of the analyses described below, we refer to the S3.

##### Second baseline: limits of agreement

Bland-Altman analyses will be performed to test for a systematic effect of repeated measurements (paired t-tests comparing the two baselines (T-1 and T0)) and to establish the limits of agreement for the outcome variables. The limits of agreement thus established will provide the basis for evaluating any effect sizes found in the main analyses.

##### Comparison to reference group

The reference data as collected in Amsterdam will be compared to the baseline data of the age- and sex-matched PwPD from the other sites. The endpoints to be compared are: the comfortable gait speed, stride length, stride time, R^2^ from the foot placement model, foot placement error (i.e. standard deviation of the residual of the foot placement model), the position gain (i.e. 𝛽_𝑝𝑜𝑠_), the velocity gain (i.e. 𝛽_𝑣𝑒𝑙_), beta power and corticomuscular coherence.

Independent t-tests will be used to test for differences between control and PD groups. For the EEG outcomes (i.e. beta power and corticomuscular coherence) cluster-based permutation tests time locked to the gait cycle will be used instead. The presence and direction of potential differences between PwPD and control can facilitate the interpretation of potential PwPD training effects.

##### Training effects of SDTT and additional value of SDTT+

To assess SDTT and SDTT+ training effects on our primary and secondary endpoints (see S3 for more detail), we will use linear mixed effect models with the interaction of treatment (SDTT, SDTT+) and time (T0,T1,T2) in the model and a random intercept for participant [79], controlling for relevant potential confounders (e.g., age, H&Y stage). Separate models will be fitted for Kiel and Sydney, whereas, as planned, the data for Bologna and Tel Aviv will be pooled. For the linear mixed effect model of the pooled data of Bologna and Tel Aviv the random intercept for participant will be nested within center [79].

To similarly assess SDTT and SDTT+ training effects in beta power and corticomuscular coherence, we will use cluster-based permutation tests time locked to the gait cycle.

##### Comparison between different types of SDTT+ interventions

The data of all participants will be used to explore the benefit of different types of SDTT+, using a linear mixed effect model with the interaction of treatment (SDTT,SDTT+VR,SDTT+MP,SDTT+VR+MP) and time (T0,T1,T2) and a random intercept for participant) and center (Tel Aviv, Bologna, Kiel, Sydney) [79], controlling for relevant potential confounders (e.g., age, H&Y stage).

##### Mechanisms underlying changes in gait performance

The data of all participants will be used to assess the mechanisms underlying improvements in gait performance. We will correlate changes in primary outcome (walking speed) with changes in biomechanical (foot placement coordination) and neural (EEG beta band power, corticomuscular coherence) outcomes. Multilevel models will be used to account for data collected at different sites.

For outcomes T2-T0, potential predictors of clinical outcomes will include independent variables with a time lag (T1-T0) and without time lag (T2-T0).

##### Mechanisms underlying changes in gait performance in daily life

To test associations between daily life and gait performance, we will correlate changes in daily-life outcomes with changes in lab-based gait performance outcomes. Multilevel models will be used to account for data collected at different sites. We will perform mediation analysis to assess the role of gait efficacy (mGES) in these changes. In addition, in an exploratory analysis we will use machine learning to generate prediction models of the daily life gait outcomes based on the lab-based outcomes and individual characteristics such as age, disease duration, and disease severity.

##### Towards understanding individual responses

Machine learning models (e.g., regularised logistic regression, decision trees, and tree-ensemble methods), aimed at identifying characteristics of responders versus non-responders, will be evaluated using nested cross-validation, with a focus on interpretability through feature-importance metrics and SHapely Additive exPlanations (SHAP) analyses. This integrative approach will allow us to determine the relative contribution of neural, biomechanical, behavioural, and experimental factors to both lab-based and daily-life gait improvements.

### Dissemination and open-access policy

The software to synchronize the different devices in the experimental setup and the analysis scripts will be made publicly available to enhance transparency, reproducibility and the potential for collaborative improvement[80].

Data will be published open access for further use of the dataset beyond the goals of the StepuP project on OpenNeuro and the Michael J. Fox Foundation (MJFF) database.

## Discussion

Treadmill training has been demonstrated to be an effective intervention for enhancing gait performance in PwPD [81]. Despite this evidence, the mechanical and neural mechanisms underlying gait improvements remain to be clarified, limiting the ability to optimize and individualize rehabilitation strategies. In this context, the StepuP consortium provides a framework to investigate these mechanisms and the factors that predict a beneficial response to treadmill training, thus moving towards a more personalized intervention. A better understanding of the mechanism may enable targeted allocation of speed dependent treadmill training to individuals most likely to benefit and could potentially facilitate the personalization of rehabilitation strategies to optimize functional outcomes.

PD gait is characterized by shorter steps and a high stride frequency [82,83], demonstrating impairments in anteroposterior (AP) foot placement control. Apart from realizing forward progression, foot placement control is the most dominant stability control mechanism during walking [23], a mechanism which is impaired in individuals with PD as compared to age-matched controls [29]. Improvements in AP foot placement control in PwPD can therefore underlie the earlier positive effects of speed dependent treadmill training on walking speed and subserve enhanced stability control. As such the StepuP project mainly focuses on AP foot placement control as a mechanistic training outcome of speed dependent treadmill training. Although this focus can be motivated from a relevance standpoint supported by the literature, it is also limiting a complete understanding of the underlying mechanisms of speed dependent treadmill training. Apart from foot placement control, other (stability) mechanisms could also improve following treadmill training, making it likely that the current study encompasses some, but not all of the underlying mechanisms through which treadmill training enhances PD gait. Moreover, apart from AP foot placement control, recent work shows that mediolateral (ML) stability-related foot placement control is also impaired in PD [29]. In general stability control is more crucial in the ML direction [84], as such, we may also expect improvements in the ML direction. After all, AP and ML stability control are not completely independent [38,85]. Our project is expected to generate a rich and comprehensive dataset, allowing for exploratory research into training effects on other (stability) mechanisms.

That the StepuP study is one of the first longitudinal clinical trials to use mobile EEG in PD adds to the novelty and richness of the to be collected dataset. Mobile EEG permits the measurement of motor-cortical beta activity (13-30 Hz) with high temporal resolution during treadmill walking to track training-induced changes in beta power and corticomuscular coherence. Beta activity of the motor cortex is strongly linked to motor control [86]. In PD, beta activity is thought to index an abnormally stable, inflexible motor state that can impair rapid adjustments of the gait cycle [33,34], which could underlie the reported imprecise foot placement control in PD [29]. Since walking generates large artefacts, rigorous pre-processing and careful interpretation of the collected EEG data is required. Independent component analysis (ICA) has been used before to successfully pre-process EEG data collected during walking [29]. Although it is unlikely that all artefacts can be removed, the quality of EEG data cleaned using ICA proved sufficient to find meaningful differences in EEG beta power during externally stabilized as compared to normal treadmill walking [32]. Since external lateral stabilization allows for diminished stability-related foot placement control [24,87] similar, yet opposite, modulations in beta power are expected following speed dependent treadmill training. As such, we are optimistic in the potential of our EEG recordings to improve the understanding of the underlying mechanisms of SDTT.

A better understanding of the underlying mechanisms is also expected to result from the SDTT+ conditions. SDTT+ is intentionally implemented as a family of destabilizing and context-enriched gait training paradigms. While all participants receive the same SDTT backbone, SDTT+ introduces additional demands – virtual obstacles (VR), mechanical perturbations (MP), or their combination. These additions may differentially affect foot placement control. Introducing MP increases the difficulty of the training by inducing larger variations in the CoM’s position and velocity. This may strengthen the coupling between pelvis movement and foot placement more than conventional treadmill training, and as such enhance training effects. Furthermore, adding visual flow through VR could engage additional feedback mechanisms, particularly those involving CoM velocity feedback [88,89]. The latter may facilitate transfer of treadmill training effects to real-world gait. Feedback of CoM velocity has been shown to play a greater role during overground walking compared to treadmill walking, likely due to the presence of visual flow [89]. Therefore, treadmill contexts closer to daily-life gait may induce more beneficial training effects. Real-world environments impose more complex and variable demands that are not reflected by standard treadmill walking. SDTT+, which elicits both proactive and reactive gait adaptations, may better reflect real-world challenges and thereby enhance functional outcomes in daily life. At the very least, the SDTT+ conditions are expected to induce a necessary variation in the training effects, facilitating the planned use of regression analyses to improve our mechanistic understanding of speed dependent treadmill training.

Although treadmill training has been shown to improve gait performance under controlled laboratory conditions, it remains uncertain whether these improvements translate to daily life. As discussed above, differences between the treadmill and real-world context may affect transfer to daily-life gait, but we may also unravel individual characteristics which can impact the training effects and their transfer. Therefore, the analysis of real-world IMU data is an important aspect of this project. Since medication status strongly influences mobility in PD and fluctuates throughout the day, we will account for medication intake timing as a potential confounding factor to better understand when and why laboratory-based training effects do or do not translate to daily-life gait. Cultural differences may also play a role, something which, to our unique advantage, can be explored within an international consortium such as StepuP.

This trial is being conducted at four sites located across three continents: Europe, Asia, and Australia. This approach enables the study to consider the diverse lifestyles of participants and the various aspects of PD associated with different geographical locations. For example, there is evidence that the presentation of dyskinesia, non-motor symptoms and comorbidities differs between PwPD in the Western Pacific Region compared to those in Europe and North America [90]. Therefore, our sample will be representative of a wide range of PD phenotypes due to the participants’ locations and environmental circumstances. Furthermore, by collecting detailed information on clinical and everyday-relevant parameters within the cohort, we will be able to conduct post-hoc analyses that address this issue and have the potential to provide new insights into PD-related disparities based on location. It should be noted however, that although the sample size is adequate for the primary clinical objective and pre-specified mixed-effects analyses, it might not be for such additional analyses, more complex mediation and machine-learning approaches. These analyses will therefore necessarily be exploratory.

### Values of the StepuP consortium

Naturally the consortium is committed to conducting an ethically responsible study. Although the burden on the participants can be considered substantial given the many lab visits and prolonged home-based assessments, participants will benefit from participation in an already proven effective [81] training intervention. As such the benefits are considered to balance, and even outweigh, the burden of participation. An important choice of our experimental design is to have a second baseline rather than a control group to be able to verify that any training effects can be contributed to the training, as opposed to potential test re-test or placebo effects. Due to the limited time frame and resources of the study we would not be able to provide the control group the full training program following the control measurements. This is considered to be unethical, as participants randomized to this group would then miss out on the effective training. By using a second baseline we have implemented an ethical solution to overcome this challenge.

In the context of large, international and multidisciplinary studies, adoption of Open Science practices is ethical imperative. Sharing setup scripts, analysis pipelines and fully annotated code fosters transparency, enhances reproducibility and enables other groups to verify, extend or adapt the work. Beyond these direct benefits, enabling data and scripts to be FAIR (findable, accessible, interoperable, reusable) is especially critical in clinical studies. By maximizing reuse of existing data and reducing redundant recruitment burdens, Open Science helps to avoid exposing additional participants to study-related burdens in the future.

Apart from ethical responsibility and Open Science, as a consortium, we value participant involvement. We expect our semi-structured interviews to reveal valuable information about the current training intervention, and important feedback to be incorporated in future research studies. Participant experiences can be used to guide future refinement of treadmill and digital adjuncts. As such they will inform iterative enhancements to training protocol, usability features, and support strategies for home-based activity.

Altogether, with this protocol, the StepuP consortium aims to understand the neuromechanical mechanisms underlying speed dependent treadmill training and its transfer to real-world walking, in order to better respond to the personal needs of individuals with PD.

## Supporting information

S1 Questionnaire for healthy participants

S2 Real-world assessment

S3 Analysis plan

S4 Spirit checklist

Approved ethical protocol

## Notes

### Competing Interest Statement

A/Prof Matthew Brodie and Dr Yoshi Okubo declare the following competing interests related to the Walking Tall mobile application used for the home-based retention training in this study. The Walking Tall App was developed by Matthew Brodie at the University of New South Wales (UNSW) in partnership with UNSW spinout Walking Tall Health and is freely available via the Apple App Store and Google Play by searching "Walking Tall". The app has been translated into multiple languages and has thousands of users globally. A/Prof Brodie is the Cofounder and Chief Executive Officer (CEO) of Walking Tall Health, the company associated with ongoing development and dissemination of the Walking Tall App. Dr Okubo is a shareholder in Walking Tall Health. These roles and interests could be perceived as a financial competing interest in relation to the use and visibility of the app within this research. All other authors declare no competing interests.

### Clinical Trial

The trial has been registered at clinicaltrials.gov (ids: NCT07057219, NCT07105787, NCT07058285, NCT06538909).

### Funding Statement

This research was financially supported by the EU Joint Programme-Neurodegenerative Disease Research (JPND), grant number JPND2022-128. The national funding agencies funding this JPND project are ZonMw, the Italian Ministry of Health, DLR Projekttrager, National Health & Medical Research Council, Israel Ministry of Health and the Swiss National Science Foundation. This study was independently designed and executed by the authors. The funding agencies were not involved in the study's design, the analysis of the results, or the final approval of the manuscript.

### Author Declarations

Ethical approval was obtained at the individual sites (Kiel D 629/24, Tel Aviv 0569-23-TLV, Sydney iRECS5114, Bologna 71-2024-SPER-AUSLBO, Amsterdam VCWE-2025-083). All participants will provide written informed consent and the research will be performed in accordance with the Declaration of Helsinki.

