## Supplementary material for "Steps against the burden of Parkinson’s disease (StepuP): Protocol of a randomized controlled trial elucidating the biomechanical and neurophysiological mechanisms of a speed dependent treadmill training intervention": S1 Questionnaire for healthy participants

*Supplementary Table 1. Inclusion-exclusion questionnaire for healthy participants in Amsterdam*

**If an answer was "yes" on any of the questions printed in "bold" participants were excluded from participation.**

| Topic | Question | No | Yes | Additional comment |
|---|---|---|---|---|
| Walking ability | Do you use a walking aid (if so, what and how often)? | | | |
| | **Is it difficult for you to walk for 7 minutes at a time without resting?** | | | |
| | **Is it difficult for you to walk for 17 minutes in one session without an aid but with breaks in between?** | | | |
| | Do you experience pain when walking? | | | |
| | Do you experience more pain when walking for longer periods? | | | |
| Risk of falling | Have you fallen in the past year? (If yes, how often)? | | | |
| | Have you fallen more than twice in the past year? | | | |
| Neurological and psychiatric disorders | **Do you have Parkinson's disease?** | | | |
| | **Do you ever experience tingling or numbness in your hands, feet, or legs (for example, neuropathy due to diabetes)?** | | | |
| | **Do you have any neurological complaints for which you have visited a neurologist in the past year (if so, what were they for)?** | | | |
| | **Do you have a psychiatric condition?** | | | |
| Medication | **Have you used sleeping pills or tranquilizers in the past week? (benzodiazepines such as oxazepam, temazepam, diazepam, zoplidem, etc.)** | | | |
| | **Have you used antidepressants in the past week?** | | | |
| | **Have you used beta-blockers in the past week?** | | | |
| | **Have you used antiepileptic drugs (such as carbamazepine, clonazepam, levetiracetam, sodium valproate, etc.) in the past week?** | | | |
| Joint disorders | **Do you suffer from osteoporosis (bone loss) or osteoarthritis (joint wear and tear)?** | | | |
| | **Do you suffer from rheumatoid arthritis (joint inflammation)?** | | | |
| | **Do you have an artificial joint (hip or knee prosthesis)?** | | | |
| Lower extremity injury | **Did you break your leg last year?** | | | |
| | **Do you suffer from rheumatoid arthritis (joint inflammation)?** | | | |
| | **Did you tear your knee or ankle ligaments last year?** | | | |
| Vestibular | **Do you often feel dizzy?** | | | |

| | |
|---|---|
| disorders Cardiovascular problems | Have you ever had a cardiac arrest and/or bypass surgery? |
| | Have you ever had chest pain? |
| | **Have you ever had stroke?** |
| | Have you ever had a pulmonary embolism? |
| | Do you have high blood pressure (systolic >140, diastolic >90) and/or are you taking medication for high blood pressure? |
| | Do you have high cholesterol (>5.2) and/or are you taking medication for high cholesterol? |
| | Have you ever fainted in the past 6 months? |
| | Do you have any other heart problems (e.g., palpitations, heart murmur, shortness of breath) for which you have visited a cardiologist in the past year (if yes, for what)? |
| Vision and hearing | Do you have trouble reading the newspaper (possibly with glasses or a magnifying glass)? |
| | Do you have trouble recognizing someone's face from a distance of 4 meters (possibly with glasses)? |
| | Do you have trouble hearing my questions clearly? |
