## Supplementary material for "Steps against the burden of Parkinson’s disease (StepuP): Protocol of a randomized controlled trial elucidating the biomechanical and neurophysiological mechanisms of a speed dependent treadmill training intervention": S2 Real-world assessment

Supplementary Table 2: Real-world assessment diary physical activity: “Please indicate every day the times when you carried out specific physical activities each of the last days in the table on the right side. These activities may include outdoor walking/hiking (W), swimming (S), guided physiotherapy (P), cycling (C), fitness (F) or dancing (D). You can specify any additional forms of physical activity that you regularly engage in, and which haven't been mentioned previously, with your investigator. Enter the corresponding abbreviation (in brackets after the respective activity) in the table. The table below serves as an example.”

| | AM | | | | | | | PM | | | | | | | | |
|---|---|---|---|---|---|---|---|---|---|---|---|---|---|---|---|---|
| Day/ Time | 6-7 | 7-8 | 8-9 | 9-10 | 10-11 | 11-12 | 12-1 | 1-2 | 2-3 | 3-4 | 4-5 | 5-6 | 6-7 | 7-8 | 8-9 | 9-10 |

Supplementary Table 3. Real-world assessment diary freezing of gait and falls: “Please indicate in the right side of the table whether you experienced freezing of gait episodes (Freezing of gait is the temporary, brief (from seconds to a few minutes), and involuntary inability to walk or to move forward. In particular, some patients with Parkinson’s disease experience the feeling of their feet being ‘glued’ to the floor. ) during the day and estimate the number of episodes during the morning ("Nr AM", before lunch) and the afternoon ("Nr PM", after lunch). If you happen to fall during this week, please also indicate the approximate time of any fall with X in the calendar”

| FOG | | | |
|---|---|---|---|
| Yes | No | Nr AM | Nr PM |
| ☐ | ☐ | | |
| ☐ | ☐ | | |
| ☐ | ☐ | | |
| ☐ | ☐ | | |
| ☐ | ☐ | | |
| ☐ | ☐ | | |
| ☐ | ☐ | | |

Supplementary Table 4: Real-world assessment diary medication intake: “Please use the table on the right side to indicate the time, (7-8, ... 9-10, as columns), for your scheduled medication. Indicate every day your medication intake. In case your actual intake matches with the scheduled intake, please indicate an X under the column “Normal day” at the relevant day. When the actual intake deviates the scheduled by more than half an hour, please indicate the actual time of intake by marking X under the relevant column for time and in the respective row with the current date. Please see the table below as an example.”

| | | AM | | | | | | | PM | | | | | | | | |
|---|---|---|---|---|---|---|---|---|---|---|---|---|---|---|---|---|---|
| | Time | 6-7 | 7-8 | 8-9 | 9-10 | 10-11 | 11-12 | 12-1 | 1-2 | 2-3 | 3-4 | 4-5 | 5-6 | 6-7 | 7-8 | 8-9 | 9-10 |
| | Scheduled Medication | | | | | | | | | | | | | | | | |
| Day | Normal Day | Actual medication intake | | | | | | | | | | | | | | | |
| | ☐ | | | | | | | | | | | | | | | | |
| | ☐ | | | | | | | | | | | | | | | | |
| | ☐ | | | | | | | | | | | | | | | | |
| | ☐ | | | | | | | | | | | | | | | | |
| | ☐ | | | | | | | | | | | | | | | | |
| | ☐ | | | | | | | | | | | | | | | | |
| | ☐ | | | | | | | | | | | | | | | | |

Supplementary Table 5. Real world questionnaire (filled once at the end of the seven days)

1. Did you use any walking aids (rollator, walking sticks, etc) during the last 7 days?
   a. Yes
   b. No
2. Was the last week normal compared to the rest of the year?
   a. Yes, the last week was a typical week.
   b. No, I did more physical activity than usual, because (please select all options that apply to you:
      - ☐ The weather allowed me to participate in physical activity
      - ☐ I felt more energized/less fatigued than usual this week
      - ☐ I was motivated to do more because of participating in this study
      - ☐ I had a new exercise routine or class that increased my physical activity
      - ☐ I experienced an improvement in health or symptoms that made it easier to be active
      - ☐ I had more social opportunities to be active (e.g., with friends, family, or groups)
      - ☐ I had more time available for physical activity this week
      - ☐ Other:

   c. No, I did less physical activity than usual, because (please select all options that apply to you):
      - ☐ The weather made it difficult to participate in physical activity
      - ☐ I felt more physically fatigued than usual this week
      - ☐ Low motivation made physical activity more challenging this week
      - ☐ I was on holiday or had visitors that disrupted my usual physical activity schedule
      - ☐ Injury made physical activity more challenging this week
      - ☐ Parkinson's symptoms made physical activity more challenging this week
      - ☐ I experienced a cold or other illness that made physical activity more challenging this week
      - ☐ Other:

3. How tired/fatigued did you feel the last days?
   Refer to the picture scale below and choose the number that best matches your mental and physical fatigue of the last seven days. Mental fatigue can make it difficult to concentrate, e.g. when reading a book or when engaging in a conversation. You might get distracted or you may also feel to be unusually irritable easily. Physical fatigue can make it difficult to perform everyday tasks, such as climbing stairs or standing for long periods. You might feel weak or unusually exhausted, even after minor efforts.

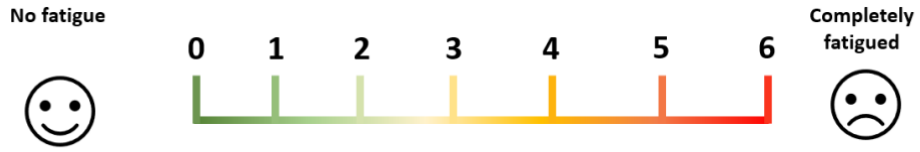
