## Supplementary material for "Steps against the burden of Parkinson’s disease (StepuP): Protocol of a randomized controlled trial elucidating the biomechanical and neurophysiological mechanisms of a speed dependent treadmill training intervention": S3 Analysis plan

**Analysis plan StepuP**

This document describes the statistical analyses to be applied to the StepuP data. In StepuP, we investigate the effects of a proven effective intervention SDTT in individuals with PD. The data set will consist of kinematic, EEG, EMG and clinical data collected in people living with PD (pts) and and age-matched reference group of individuals without PD (ref) at different centers. The total sample is composed as follows**:**

| | pts | pts | pts | pts | ref | |
|---|---|---|---|---|---|---|
| | 1. SDTT | 2. SDTT+VR | 3. SDTT+MP | 4. SDTT+VR+MP | | Total participants |
| Tel Aviv | 11 | 10 | | | | 21 |
| Bologna | 10 | 11 | | | | 21 |
| Kiel | 21 | | 21 | | | 42 |
| Sydney | 21 | | | 21 | | 42 |
| Amsterdam | | | | | 21 | 21 |

Our main aim is to determine the mechanisms underlying effects of SDTT and SDTT+ on gait performance in clinical tests and in daily-life. This entails an analysis of pooled data (excluding "ref"). Nested within this design, several randomized comparisons will be made to assess the added value of the SDTT+ interventions over SDTT. To interpret any effects in the absence of a placebo control group, it is important to determine the minimal meaningful changes for the outcome parameters. In addition, a comparison between participants with and without PD will be done to support interpretation of effects in the PD group. The analyses to this end are described as analyses 1a and 1b. Subsequently the nested randomized comparisons are described as analyses 2a to 2c. In analysis 2d we will compare the different treatment protocols. Finally, the analysis to test hypotheses on the causality of changes in gait performance are described as analyses 3a and 3b.

**Analysis 1a: reliability**

**Population:**
From the populations measured in Sydney,  participants chosen at random will participate in an additional measurement set 4 weeks prior to start of the intervention (T-1). Sydney will aim for 21 participants to be enrolled in T-1, or a minimum of 16 participants, if practical constraints prevent to reach the full sample. The final sample is kept small for pragmatic reasons. First, it is not feasible to apply this additional measurement in all participants at all centers. Second, in case a systematic effect of repeated testing is found, this may need to be statistically corrected for or the sub-sample of individual participants with this additional baseline may need to omitted in the pooled analyses (3a and 3b).

| | | |
|---|---|---|
| Center | T-1 | T0 |
| Sydney | 21 (/16) | 21 (/16) |

**Dependent variables:**
primary clinical outcomes:
- preferred gait speed (20 m walk test)
- stride length (at preferred speed)
- stride length (at 1.4 km/h)

secondary clinical outcomes:
- stride length variability (at preferred speed and at 1.4 km/h)
- stride time (at preferred speed and at 1.4 km/h)
- stride time variability (at preferred speed and at 1.4 km/h)
- step width (at preferred speed and at 1.4 km/h)
- step width variability (at preferred speed and at 1.4 km/h)
- falls (diary)
- timed-up-and-go (TUG) test
- 2-minute walk test
- MiniBESTest
- MOCA
- MDS-UPDRS, part III
- mGES

feedback control variables (all at 1.4 km/h):
- $R^2$ (feedback model relating foot placement to centre of mass movement)
- RMS error (feedback model relating foot placement to centre of mass movement)
- Position gain (feedback model relating foot placement to centre of mass movement)
- Velocity gain (feedback model relating foot placement to centre of mass movement)

neural correlates (T1 all at 1.4 km/h):
- EEG beta power in(pre-)motor areas during walking at preferred speed
- EEG-EMG coherence during walking at preferred speed

**Analysis technique:**
Bland-Altman analyses will be performed to test for a systematic effect of repeated measurements (paired t-tests) and to establish the limits of agreement for the outcome variables. The limits of agreement thus established will provide the basis for evaluating any effect sizes found in analyses 2a-2c.

**Analysis 1b: case-control study**

**Population:**
21 (min. 17 for analysis) individuals with PD (T0) randomly selected from total sample
21 (min. 17 for analysis) healthy individuals age- and sex-matched to PD group

**Independent variables:**
group (PD, control)

**Dependent variables:**
primary clinical outcomes:

- preferred gait speed (20 m walk test)
- stride length (at preferred speed and at 1.4 km/h)

secondary clinical outcomes:

- stride length variability (at preferred speed and at 1.4 km/h)
- stride time (at preferred speed and at 1.4 km/h)
- stride time variability (at preferred speed and at 1.4 km/h)
- step width (at preferred speed and at 1.4 km/h)
- step width variability (at preferred speed and at 1.4 km/h)

feedback control variables (at 1.4 km/h):

- $R^2$ (feedback model relating foot placement to centre of mass movement)
- RMS error (feedback model relating foot placement to centre of mass movement)
- Position gain (feedback model relating foot placement to centre of mass movement)
- Velocity gain (feedback model relating foot placement to centre of mass movement)

neural correlates (at 1.4 km/h):

- beta power during walking at preferred speed
- EEG-EMG coherence during walking at preferred speed

**Analysis technique:**
Initially, independent t-tests and for EEG data cluster-based permutation tests time locked to the gait cycle will be used to compare individuals with PD to controls. The presence of and direction of differences between groups will facilitate interpretation for any effects found in analyses 2a - 2c. For this no corrections for multiple comparisons will be made. However, to present overall differences between groups we will reduce dimensionality of the set of primary and secondary clinical outcomes by means of PCA and use stepwise logistic regression on PC-scores to assess between group differences. For feedback control variables and neural correlates we will separately correct for multiple comparisons.

**Analysis 2a: RCT SDTT vs SDTT+**

Added value of additional component (SDTT+) to Speed Dependent Treadmill Training (SDTT)

**Population:**

| | pts | pts | pts |
|---|---|---|---|
| | SDTT | SDTT+VR | Total |
| Tel Aviv | 11 | 10 | 21 |
| Bologna | 10 | 11 | 21 |

**Independent variables:**
time (T0,T1,T2) x group (SDTT, SDTT+)

**Dependent variables:**
primary clinical outcomes:

- preferred gait speed (20 m walk test)
- stride length (at preferred speed)
- stride length (at 1.4 km/h)

secondary clinical outcomes (T1):

- stride length variability (at preferred speed and at 1.4 km/h)
- stride time (at preferred speed and at 1.4 km/h)
- stride time variability (at preferred speed and at 1.4 km/h)
- step width (at preferred speed and at 1.4 km/h)
- step width variability (at preferred speed and at 1.4 km/h)
- falls (diary)
- timed-up-and-go (TUG) test
- 2-minute walk test
- MiniBESTest
- MOCA
- MDS-UPDRS, part III
- mGES

secondary clinical outcomes (T2):

- steps per day
- uninterrupted walk durations
- stride time variability
- symmetry

feedback control variables (T1 all at 1.4 km/h):

- $R^2$ (feedback model relating foot placement to centre of mass movement)
- RMS error (feedback model relating foot placement to centre of mass movement)
- Position gain (feedback model relating foot placement to centre of mass movement)
- Velocity gain (feedback model relating foot placement to centre of mass movement)

neural correlates (T1 all at 1.4 km/h):

- EEG beta power in(pre-)motor areas during walking at preferred speed
- EEG-EMG coherence during walking at preferred speed

**Analysis technique:**
Linear mixed effect models with the interaction of treatment and time in the model and a random intercept for participant nested within center (Twisk et al., 2018), controlling for relevant potential confounders (e.g., age, H&Y stage). For the EEG data, we will use cluster-based permutation tests time locked to the gait cycle.

**Analysis 2b and 2c: 2 independent RCTs SDTT vs SDTT+**

Added value of additional component (SDTT+) to Speed Dependent Treadmill Training (SDTT)

**Population:**

| | pts | pts | pts | pts | pts |
|---|---|---|---|---|---|
| | 1. SDTT | | 3. SDTT+MP | 4. SDTT+VR+MP | Total participants |
| Kiel | 21 | | 21 | | 42 |
| Sydney | 21 | | | 21 | 42 |

**Independent variables:**
time (T0,T1,T2) x group (SDTT, SDTT+)

**Dependent variables:**
primary clinical outcomes:

- preferred gait speed (20 m walk test)
- stride length (at preferred speed)
- stride length (at 1.4 km/h)

secondary clinical outcomes (T1):

- stride length variability (at preferred speed and at 1.4 km/h)
- stride time (at preferred speed and at 1.4 km/h)
- stride time variability (at preferred speed and at 1.4 km/h)
- step width (at preferred speed and at 1.4 km/h)
- step width variability (at preferred speed and at 1.4 km/h)
- falls (diary)
- timed-up-and-go (TUG) test
- 2-minute walk test
- MiniBESTest
- MOCA
- MDS-UPDRS, part III
- mGES

secondary clinical outcomes (T2):

- steps per day
- uninterrupted walk durations
- stride time variability
- symmetry

feedback control variables (T1 all at 1.4 km/h):

- $R^2$ (feedback model relating foot placement to centre of mass movement)
- RMS error (feedback model relating foot placement to centre of mass movement)
- Position gain (feedback model relating foot placement to centre of mass movement)
- Velocity gain (feedback model relating foot placement to centre of mass movement)

neural correlates (T1 all at 1.4 km/h):

- EEG beta power in(pre-)motor areas during walking at preferred speed
- EEG-EMG coherence during walking at preferred speed

**Analysis technique:**
Linear mixed effect models with the interaction of treatment and time in the model and a random intercept for participant (Twisk et al., 2018), controlling for relevant potential confounders (e.g., age, H&Y stage). For the EEG data, we will use cluster-based permutation tests time locked to the gait cycle.

**Analysis 2d: Pooled RCT SDTT vs SDTT+**

### Comparison of the different SDTT+ interventions

**Population:**

| | pts | pts | pts | pts | pts |
|---|---|---|---|---|---|
| | 1. SDTT | 2. SDTT+VR | 3. SDTT+MP | 4. SDTT+VR+MP | Total participants |
| Tel Aviv | 11 | 10 | | | 21 |
| Bologna | 10 | 11 | | | 21 |
| Kiel | 21 | | 21 | | 42 |
| Sydney | 21 | | | 21 | 42 |

**Independent variables:**
time (T0,T1,T2) x group (SDTT, SDTT+VR,SDTT+MP,SDTT+VR+MP)

**Dependent variables:**
primary clinical outcomes:

- preferred gait speed (20 m walk test)
- stride length (at preferred speed)
- stride length (at 1.4 km/h)

secondary clinical outcomes (T1):

- stride length variability (at preferred speed and at 1.4 km/h)
- stride time (at preferred speed and at 1.4 km/h)
- stride time variability (at preferred speed and at 1.4 km/h)
- step width (at preferred speed and at 1.4 km/h)
- step width variability (at preferred speed and at 1.4 km/h)
- falls (diary)
- timed-up-and-go (TUG) test
- 2-minute walk test
- MiniBESTest
- MOCA
- MDS-UPDRS, part III
- mGES

secondary clinical outcomes (T2):

- steps per day
- uninterrupted walk durations
- stride time variability
- symmetry

feedback control variables (T1 all at 1.4 km/h):

- $R^2$ (feedback model relating foot placement to centre of mass movement)
- RMS error (feedback model relating foot placement to centre of mass movement)
- Position gain (feedback model relating foot placement to centre of mass movement)
- Velocity gain (feedback model relating foot placement to centre of mass movement)

neural correlates (T1 all at 1.4 km/h):

- EEG beta power in(pre-)motor areas during walking at preferred speed
- EEG-EMG coherence during walking at preferred speed

**Analysis technique:** Linear mixed effect models with the interaction of treatment and time in the model, a random intercept for center, and a random intercept for participant (Twisk et al., 2018), controlling for relevant potential confounders (e.g., age, H&Y stage). For the EEG data, we will use cluster-based permutation tests time locked to the gait cycle.

**Analysis 3a: Mechanisms underlying changes in gait performance**

**Population:**
124 individuals with PD

Associations between clinical outcomes and feedback control

**Dependent variables:**
primary clinical outcomes (T1-T0 / T2-T0):

- ∆ preferred gait speed (20 m walk test)
- ∆ stride length (at preferred speed and at 1.4 km/h)

secondary clinical outcomes (T1-T0 / T2-T0):

- ∆ stride length variability (at preferred speed and at 1.4 km/h)
- ∆ stride time (at preferred speed and at 1.4 km/h)
- ∆ stride time variability (at preferred speed and at 1.4 km/h)
- ∆ step width (at preferred speed and at 1.4 km/h)
- ∆ step width variability (at preferred speed and at 1.4 km/h)

**Independent variables:**
feedback control variables (T1-T0 / T2-T0, at 1.4 km/h):

- ∆ $R^2$ (feedback model relating foot placement to centre of mass movement)
- ∆ RMS error (feedback model relating foot placement to centre of mass movement)
- ∆ Position gain (feedback model relating foot placement to centre of mass movement)
- ∆ Velocity gain (feedback model relating foot placement to centre of mass movement)

neural correlates (T1-T0 / T2-T0, at 1.4 km/h):

- ∆ beta power during walking at preferred speed
- ∆ EEG-EMG coherence during walking at preferred speed

Associations between feedback control variables and neurological correlates

**Dependent variables:**
feedback control variables (T1-T0 / T2-T0, at 1.4 km/h):

- ∆ $R^2$ (feedback model relating foot placement to centre of mass movement)
- ∆ RMS error (feedback model relating foot placement to centre of mass movement)
- ∆ Position gain (feedback model relating foot placement to centre of mass movement)
- ∆ Velocity gain (feedback model relating foot placement to centre of mass movement)

**Independent variables:**
neural correlates (T1-T0 / T2-T0, at 1.4 km/h):

- ∆ beta power during walking at preferred speed
- ∆ EEG-EMG coherence during walking at preferred speed

**Analysis technique:**
We will correlate changes in primary outcomes (e.g., walking speed) with changes in behavioral (foot placement coordination) and neural (EEG beta band power, cortico-muscular coherence) outcomes. Multilevel models will be used, to account for data collected at different sites. For outcomes T2-T0, potential predictors of clinical outcomes will include independent variables with a time lag (T1-T0) and without time lag (T2-T0).

**Analysis 3b: Mechanisms underlying changes in gait performance in daily life**

**Population:**
124 individuals with PD

Associations between daily life gait and gait performance

**Dependent variables:**
secondary clinical outcomes (T2-T0):

- Δ steps per day
- Δ uninterrupted walk durations
- Δ stride time variability
- Δ symmetry

**Independent variables:**
primary clinical outcomes (T2-T0):

- Δ preferred gait speed (20 m walk test)
- Δ stride length (at preferred speed)

secondary clinical outcomes (T2-T0):

- Δ stride length variability (at preferred speed)
- Δ stride time (at preferred speed)
- Δ stride time variability (at preferred speed)
- Δ step width (at preferred speed)
- Δ step width variability (at preferred speed)

feedback control variables (T2-T0):

- Δ $R^2$ (feedback model relating foot placement to centre of mass movement)
- Δ RMS error (feedback model relating foot placement to centre of mass movement)
- Δ Position gain (feedback model relating foot placement to centre of mass movement)
- Δ Velocity gain (feedback model relating foot placement to centre of mass movement)

gait efficacy (T2-T0):

- Δ mGES

**Analysis technique:**
We will correlate changes in daily-life outcomes with changes in lab-based gait performance outcomes. Multilevel models will be used to account for data collected at different sites. We will perform mediation analysis to assess the role of gait efficacy. In addition, in an exploratory analysis we will use machine learning to generate prediction models of the dependent variables listed here based on the independent variables listed and individual characteristics such as age, disease duration, and disease severity.
