## Supplementary material for "Steps against the burden of Parkinson’s disease (StepuP): Protocol of a randomized controlled trial elucidating the biomechanical and neurophysiological mechanisms of a speed dependent treadmill training intervention": S4 Spirit checklist

#### SPIRIT 2025 checklist of items to address in a randomized trial protocol*

| Section / Topic | No | SPIRIT 2025 checklist item description | Reported on page no. |
|---|---|---|---|
| **Administrative information** | | | |
| Title and structured summary | 1a | Title stating the trial design, population, and interventions, with identification as a protocol | 1 |
| | 1b | Structured summary of trial design and methods, including items from the World Health Organization Trial Registration Data Set | 5-6 |
| Protocol version | 2 | Version date and identifier | 1 |
| Roles and responsibilities | 3a | Names, affiliations, and roles of protocol contributors | 1 |
| | 3b | Name and contact information for the trial sponsor | 6 |
| | 3c | Role of trial sponsor and funders in design, conduct, analysis, and reporting of trial; including any authority over these activities | 23 |
| | 3d | Composition, roles, and responsibilities of the coordinating site, steering committee, endpoint adjudication committee, data management team, and other individuals or groups overseeing the trial, if applicable | 7 |
| **Open science** | | | |
| Trial registration | 4 | Name of trial registry, identifying number (with URL), and date of registration. If not yet registered, name of intended registry | 6 |
| Protocol and statistical analysis plan | 5 | Where the trial protocol and statistical analysis plan can be accessed | 6 |
| Data sharing | 6 | Where and how the individual de-identified participant data (including data dictionary), statistical code, and any other materials will be accessible | 15 |
| Funding and conflicts of interest | 7a | Sources of funding and other support (e.g., supply of drugs) | 6 |
| | 7b | Financial and other conflicts of interest for principal investigators and steering committee members | 23 |
| Dissemination policy | 8 | Plans to communicate trial results to participants, healthcare professionals, the public, and other relevant groups (e.g., reporting in trial registry, plain language summary, publication) | 20 |
| **Introduction** | | | |
| Background and rationale | 9a | Scientific background and rationale, including summary of relevant studies (published and unpublished) examining benefits and harms for each intervention | 2 |
| | 9b | Explanation for choice of comparator | 3 |
| Objectives | 10 | Specific objectives related to benefits and harms | 20 |
| **Methods: Patient and public involvement, trial design** | | | |
| Patient and public involvement | 11 | Details of, or plans for, patient or public involvement in the design, conduct, and reporting of the trial | 20 |
| Trial design | 12 | Description of trial design including type of trial (e.g., parallel group, crossover), allocation ratio, and framework (e.g., superiority, equivalence, non-inferiority, exploratory) | 7 |

| | | | |
|---|---|---|---|
| **Methods: Participants, interventions, and outcomes** | | | |
| Trial setting | 13 | Settings (e.g., community, hospital) and locations (e.g., countries, sites) where the trial will be conducted | 7 |
| Eligibility criteria | 14a | Eligibility criteria for participants | 7 |
| | 14b | If applicable, eligibility criteria for sites and for individuals who will deliver the interventions (e.g., surgeons, physiotherapists) | 7 |
| Intervention and comparator | 15a | Intervention and comparator with sufficient details to allow replication including how, when, and by whom they will be administered. If relevant, where additional materials describing the intervention and comparator (e.g., intervention manual) can be accessed | 13 |
| | 15b | Criteria for discontinuing or modifying allocated intervention/comparator for a trial participant (e.g., drug dose change in response to harms, participant request, or improving/worsening disease) | 13 |
| | 15c | Strategies to improve adherence to intervention/comparator protocols, if applicable, and any procedures for monitoring adherence (e.g., drug tablet return, sessions attended) | 12 |
| | 15d | Concomitant care that is permitted or prohibited during the trial | - |
| Outcomes | 16 | Primary and secondary outcomes, including the specific measurement variable (e.g., systolic blood pressure), analysis metric (e.g., change from baseline, final value, time to event), method of aggregation (e.g., median, proportion), and time point for each outcome | 18 |
| Harms | 17 | How harms are defined and will be assessed (e.g., systematically, non-systematically) | - |
| Participant timeline | 18 | Time schedule of enrollment, interventions (including any run-ins and washouts), assessments, and visits for participants. A schematic diagram is highly recommended (see Figure) | 13 |
| Sample size | 19 | How sample size was determined, including all assumptions supporting the sample size calculation | 20 |
| Recruitment | 20 | Strategies for achieving adequate participant enrollment to reach target sample size | 20 |
| **Methods: Assignment of interventions** | | | |
| Randomization: | | | |
| Sequence generation | 21a | Who will generate the random allocation sequence and the method used | 13 |
| | 21b | Type of randomization (simple or restricted) and details of any factors for stratification. To reduce predictability of a random sequence, other details of any planned restriction (e.g., blocking) should be provided in a separate document that is unavailable to those who enroll participants or assign interventions | 13 |
| Allocation concealment mechanism | 22 | Mechanism used to implement the random allocation sequence (e.g., central computer/telephone; sequentially numbered, opaque, sealed containers), describing any steps to conceal the sequence until interventions are assigned | 13 |
| Implementation | 23 | Whether the personnel who will enroll and those who will assign participants to the interventions will have access to the random allocation sequence | 13 |
| Blinding | 24a | Who will be blinded after assignment to interventions (e.g., participants, care providers, outcome assessors, data analysts) | 13 |

| | | | |
|---|---|---|---|
| | 24b | If blinded, how blinding will be achieved and description of the similarity of interventions | 13 |
| | 24c | If blinded, circumstances under which unblinding is permissible, and procedure for revealing a participant's allocated intervention during the trial | 13 |
| **Methods: Data collection, management, and analysis** | | | |
| Data collection methods | 25a | Plans for assessment and collection of trial data, including any related processes to promote data quality (e.g., duplicate measurements, training of assessors) and a description of trial instruments (e.g., questionnaires, laboratory tests) along with their reliability and validity, if known. Reference to where data collection forms can be accessed, if not in the protocol | 15 |
| | 25b | Plans to promote participant retention and complete follow-up, including list of any outcome data to be collected for participants who discontinue or deviate from intervention protocols | 20 |
| Data management | 26 | Plans for data entry, coding, security, and storage, including any related processes to promote data quality (e.g., double data entry; range checks for data values). Reference to where details of data management procedures can be accessed, if not in the protocol | 15 |
| Statistical methods | 27a | Statistical methods used to compare groups for primary and secondary outcomes, including harms | 18 |
| | 27b | Definition of who will be included in each analysis (e.g., all randomized participants), and in which group | 7 |
| | 27c | How missing data will be handled in the analysis | 18 |
| | 27d | Methods for any additional analyses (e.g., subgroup and sensitivity analyses) | 18 |
| **Methods: Monitoring** | | | |
| Data monitoring committee | 28a | Composition of data monitoring committee (DMC); summary of its role and reporting structure; statement of whether it is independent from the sponsor and funder; conflicts of interest and reference to where further details about its charter can be found, if not in the protocol. Alternatively, an explanation of why a DMC is not needed | - |
| | 28b | Explanation of any interim analyses and stopping guidelines, including who will have access to these interim results and make the final decision to terminate the trial | - |
| Trial monitoring | 29 | Frequency and procedures for monitoring trial conduct. If there is no monitoring, give explanation | - |
| **Ethics** | | | |
| Research ethics approval | 30 | Plans for seeking research ethics committee/institutional review board approval | 6 |
| Protocol amendments | 31 | Plans for communicating important protocol modifications to relevant parties | - |
| Consent or assent | 32a | Who will obtain informed consent or assent from potential trial participants or authorized proxies, and how | 6 |
| | 32b | Additional consent provisions for collection and use of participant data and biological specimens in ancillary studies, if applicable | 6 |
| Confidentiality | 33 | How personal information about potential and enrolled participants will be collected, shared, and maintained in order to protect confidentiality before, during, and after the trial | - |
| Ancillary and post-trial care | 34 | Provisions, if any, for ancillary and post-trial care, and for compensation to those who suffer harm from trial participation | - |

*We strongly recommend reading this checklist in conjunction with the SPIRIT 2025 Explanation and Elaboration and the SPIRIT 2025 Expanded Checklist for important clarifications on all the items. We also recommend reading relevant SPIRIT extensions. See www.consort-spirit.org

Citation: Chan A-W, Boutron I, Hopewell S, Moher D, Schulz KF, et al. SPIRIT 2025 statement: updated guideline for protocols of randomised trials. BMJ 2025;389:e081477. https://dx.doi.org/10.1136/bmj-2024-081477

© 2025 Chan A-W et al. This is an Open Access article distributed under the terms of the Creative Commons Attribution License (https://creativecommons.org/licenses/by/4.0/), which permits unrestricted use, distribution, and reproduction in any medium, provided the original work is properly cited.

### Steps against the burden of Parkinson's disease (StepuP): Protocol of a randomized controlled trial elucidating the biomechanical and neurophysiological mechanisms of a speed dependent treadmill training intervention

Anina Moira van Leeuwen[1,*,+], Julius Welzel[2,+], Ilaria D'Ascanio[3], Charlotte Lang[4], Vaishali Vinod[2], Pia Görrisen[2], Johanna Geritz[2], Clint Hansen[2], Eran Gazit[5], Shahar Siman Tov[5], Revital Prusak[5], Ilaria Casadei[6], Angela Contri[6], Francesca Tampellini[6], Leonardo Pellicciari[6], Giovanna Lopane[6], Giovanna Calandra Buonaura[6], Luca Palmerini[3], Noman Zahid[8], Mayna Ratanapongleka[8], Husna Razee[8], Frederic von Wegner[8,9], Bernadette C.M. van Wijk[1],[7] Sjoerd M. Bruijn[1], Deepak K. Ravi[4], Yoshiro Okubo[8,9], Navrag B. Singh[4,10] , Matthew Brodie[8], Fabio La Porta[6], Jeffrey M. Hausdorff[5,11,12], Walter Maetzler[2] and Jaap H. van Dieën[1]

[1] Department of Human Movement Sciences, Vrije Universiteit Amsterdam, Amsterdam Movement Sciences, Amsterdam, Netherlands

[2]Department of Neurology, University Hospital Schleswig-Holstein and Kiel University, Kiel, Germany

[3]Department of Electrical, Electronic, and Information Engineering "Guglielmo Marconi", University of Bologna, Bologna, Italy

[4]Institute for Biomechanics, ETH Zürich, Zürich, Switzerland

[5]Center for the Study of Movement, Cognition, and Mobility, Neurological Institute, Tel Aviv Sourasky Medical Center, Israel

[6]IRCCS Istituto delle Scienze Neurologiche di Bologna, Bologna, Italy

[7]Department of Neurology, Amsterdam University Medical Center, Amsterdam Neuroscience, University of Amsterdam, Amsterdam, the Netherlands.

[8]University of New South Wales, Sydney, Australia

[9]Neuroscience Research Australia, Sydney, Australia

[10]Future Health Technologies Programme, Singapore-ETH Centre, Singapore, Singapore

[11]Department of Physical Therapy, Gray Faculty of Medical & Health Sciences & Sagol School of Neuroscience, Tel Aviv University

[12]Rush Alzheimer's Disease Center, Rush University Medical Center and Department of Orthopaedic Surgery, Rush Medical College

*Corresponding author


+Shared first authors

Version 4.0: 04.02.2026

| | TRIAL PERIOD | | | |
|---|---|---|---|---|
| | Enrollment | | Post-randomization | Close-out |
| **TIMEPOINT** | ***-t<sub></sub>*** | | | |

| **TIMEPOINT** | **$-t_i$ to 0** | **T0** | ***T1*** | ***T2*** |
|---|---|---|---|---|
| **ENROLLMENT:** | | | | |
| **Eligibility screen** | X | | | |
| **Informed consent** | X | | | |
| **Randomization** | | X | | |
| **INTERVENTION/COMPARATOR:** | | | | |
| ***SDTT+VR, SDTT+P, SDTT+VP+R*** | | | | |
| ***SDTT (PD)*** | | | | |
| ***SDTT (healthy)*** | | X | | |
| ***Home-based training*** | | | | |
| **ASSESSMENTS/OUTCOMES:** | | | | |
| ***Clinical assessment*** | | X | X | X |
| ***Functional assessment*** | | X | X | X |
| ***Real-world assessment*** | | X | X | X |
| ***20 meter walking test*** | | X | X | X |
| ***MDS-UPDRS part III*** | | X | X | X |
| ***MiniBEST-Test*** | | X | X | X |
| ***mGES*** | | X | X | X |
| ***ShortFES-I*** | | X | X | X |
| ***EQ-5D*** | | X | X | X |

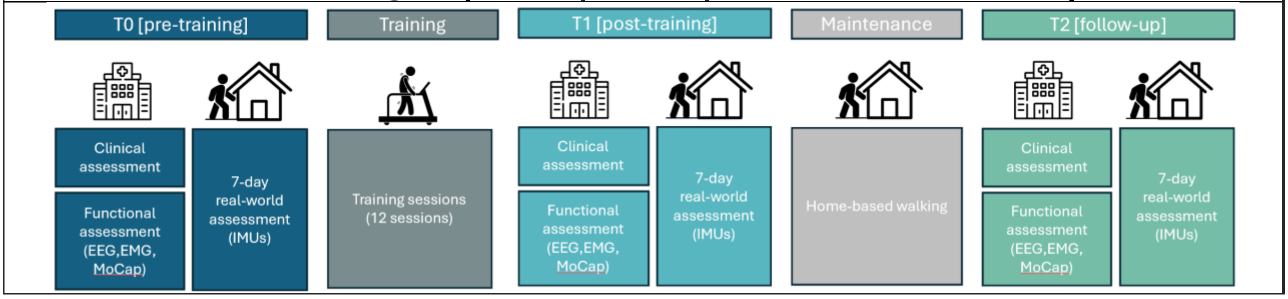


***Fig 1. Participant timeline: schedule of enrolment, intervention, and assessments.*** *EEG, elecroencephalogram; EMG, electromyography; FACIT-F, Functional Assessment of Chronic Illness Therapy–Fatigue; FES-I, Falls Efficacy Scale–International; IMU, Inertial Measurement Unit; MDS-UPDRS, Movement Disorder Society-sponsored Unified Parkinson's disease Rating Scale; mGES, modified Gait Efficacy Scale; MiniBEST, Mini-Balance Evaluation Systems Test; MoCap, motion capture.*

#### Ethical approval

Ethical approval was obtained at the individual sites (Kiel D 629/24, Tel Aviv 0569-23-TLV, Sydney iRECS5114, Bologna 71-2024-SPER-AUSLBO, Amsterdam VCWE-2025-083). All participants will provide written informed consent and the research will be performed in accordance with the Declaration of Helsinki. The trial has been registered at clinicaltrials.gov (ids: NCT07057219, NCT07105787, NCT07058285, NCT06538909).

### Participants

Participants will be enrolled in the study across the four clinical centers in accordance with the inclusion and exclusion criteria detailed in Table 1. Recruitment will be conducted via registries and hospital records or via flyers and local advertisements.

For the healthy reference group in Amsterdam, a specific exclusion criteria questionnaire (S1) will be applied, ensuring that all participants in this group do not have any (neurological) impairments or injuries which could affect their walking pattern.

***Table 1. Inclusion and exclusion criteria for participation in the StepuP trial.***

| |
|---|
| **Inclusion criteria:**<br>- Diagnosis of Parkinson's disease according to the Movement Disorder Society (MDS) criteria [48]<br>- Hoehn and Yahr (H&Y) stages I to III<br>- MDS-UPDRS gait subscore ≥1 |
| **Exclusion criteria:**<br>- Any known general health condition likely to interfere with or to pose a contraindication to non-medically supervised physical exercise (e.g., severe heart disease, severe chronic artery disease, orthostatic hypotension according to guidelines, chronic respiratory problems, subjective or objective vertigo, relevant orthopedic and other diseases such as recent lower limbs fractures limiting weight bearing, etc.)<br>- Moderate or severe depression (BDI-II ≥18)<br>- Cognitive impairment which may preclude the possibility to provide a fully informed consent to enrolment<br>- Linguistic comprehension capacity less than 75% in ordinary conversation due to, for instance, an aphasic impairment or to severe deafness notwithstanding the use of an acoustic aid, as judged by the investigator<br>- Severe psychiatric comorbidity which may interfere with compliance with the study protocol (e.g., a known diagnosis of personality disorder, documented previous occurrences of delirium and psychomotor agitation)<br>- History of or current status of substance dependency<br>- Implanted Deep Brain Stimulation device |

*Legend: BDI Beck Depression Inventory, MDS – Movement Disorder Society, PD-Parkinson's disease, UPDRS – Unified Parkinson's Disease Rating Scale.*

#### Randomization procedure

A researcher not located at a site where recordings of PwPD are performed (M.v.L. in Amsterdam) will prepare randomization sequences using a custom Python script with block randomization stratified by site and H&Y stage and securely uploaded to REDCap. The block sizes will randomly range between 2 and 4 for each of Bologna and Tel Aviv, and 2 and 6 for each of Kiel and Sydney. To ensure allocation concealment, participants will be randomized into the SDTT or SDTT+ group after the T0 assessment. This ensures that neither recruiters nor participants will be aware of group assignments beforehand, thereby minimizing the risk of selection bias.

### Intervention and control

All participants will undertake a treadmill-based intervention, comprising 12 sessions of 30 minutes each.

To understand the effects of additional components, each center will conduct SDTT and SDTT+. Tel Aviv and Bologna will apply virtual reality (VR), while Kiel will administer mechanical perturbations, and Sydney will apply both VR and mechanical perturbations (Table 2). The SDTT+VR program performed in Bologna and Tel Aviv will use the Gait Better system [21], where the participant is required to step over obstacles which appear on the screen eliciting proactive gait adaptations. The SDTT+ perturbations program performed in Kiel will employ accelerations and decelerations of the treadmill belt to elicit reactive gait adaptations. The SDTT+ program in Sydney involves both VR and perturbations where the participant is required to step over virtual obstacles and treadmill belt accelerations or decelerations occur when obstacle-foot collision occur [49], eliciting both proactive and reactive gait adaptations.

***Table 2. Intervention and Control groups.***

| Center | Intervention Group | | Control Group | |
|---|---|---|---|---|
| | **Treatment** | **N. participants** | **Treatment** | **N. participants** |
| Bologna | SDTT+VR | 11 | SDTT | 10 |
| Tel Aviv | SDTT+VR | 10 | SDTT | 11 |
| Kiel | SDTT+P | 21 | SDTT | 21 |
| Sydney | SDTT+VR+P | 21 | SDTT | 21 |

*Legend: P perturbations, SDTT Speed dependent treadmill training, VR virtual reality.*

#### Treadmill training protocol

At the first training of each training week, a single 20-meter walk test will be performed before the treadmill training in order to define the comfortable walking speed for training (CST). The CST will be used to individually tailor treadmill training intensity, ensuring that the intervention is adapted weekly to the current functional capacity of each participant.

Each training session consists of three training blocks at three different speeds (V1, V2 and V3), intermingled with recovery trials. V1 will always be set at 80% of CST, whereas V2 and V3 may progress from 80% to 110% of CST according to the training progression protocol (Table 3). Each session begins with a warm-up period (walking at 70% of CST). Between training blocks is a recovery phase where participants walk at 70% of CST for 2.5 minutes.

During training, participants will wear an overhead safety harness to prevent falls. Treadmill railings may be held only if needed and only during the first minute of each training block to allow the participant to adjust to the novel speed demands. Then, participants will be encouraged to use reciprocal arm swing.

All training sessions will be conducted individually to ensure safety and to allow close supervision of each participant. A physiotherapist will be present to monitor the participant's gait pattern and signs of fatigue but will not give any physical assistance during the walking. Furthermore, the physiotherapist will ensure that adequate rest breaks are inserted to maintain participant safety and will record their frequency and duration.

*Table 3. Progression of training speed and duration.*

| Training session # | CST recorded in session # | V1 | V2 | V3 | Training trial duration progression | Max training trial duration |
|---|---|---|---|---|---|---|
| 1 | 1 | 80% CST | 85% CST | 90% CST | 5 min* | 5 min |
| 2 | 1 | 80% CST | 85% CST | 90% CST | 30 sec | 5 min 30 sec |
| 3 | 1 | 80% CST | 90% CST | 95% CST | = | 5 min 30 sec |
| 4 | 4 | 80% CST | 90% CST | 95% CST | +30 sec | 6 min |
| 5 | 4 | 80% CST | 95% CST | 100% CST | = | 6 min |
| 6 | 4 | 80% CST | 95% CST | 100% CST | +30 sec | 6 min 30 sec |
| 7 | 7 | 80% CST | 100% CST | 105% CST | = | 6 min 30 sec |
| 8 | 7 | 80% CST | 100% CST | 105% CST | +30 sec | 7 min |
| 9 | 7 | 80% CST | 100% CST | 110% CST | = | 7 min |
| 10 | 10 | 80% CST | 100% CST | 105% CST | +30 sec | 7 min 30 sec |
| 11 | 10 | 80% CST | 100% CST | 105% CST | = | 7 min 30 sec |
| 12 | 10 | 80% CST | 100% CST | 110% CST | +30 sec | 8 min |

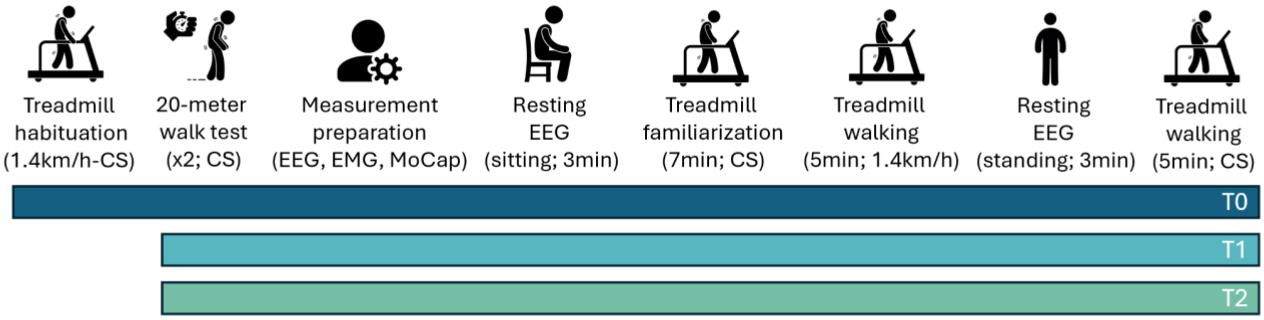

***Fig 2. Functional assessment timeline.*** *The tests performed at T0,T1, and T2 are indicated by the blue, light blue, and green lines, respectively, shown at the bottom of the figure. For each recorded task, additional information is reported in parentheses (e.g., duration, standing/sitting condition for static tasks, and velocity for dynamic tasks).*

***Table 4. Placement of motion capture markers.***

| **Marker** | **Location** |
|---|---|
| Heel# | Calcaneus |
| Pelvis# | In between the two posterior superior iliac spines |
| Asymmetric foot marker# | An additional marker at any visible location on the right foot, to ensure left and right asymmetry |
| Thigh* | Right: Over the upper 1/3 lateral surface of the thigh<br>Left: Over the lower lateral 1/3 surface of the thigh |
| Knee | On the flexion extension axis (lateral) |
| Tibia* | Right: Over the upper 1/3 surface of the right shank<br>Left: Over the lower 1/3 surface of the left shank |
| Ankle | On the lateral malleolus |
| Toe | Over the second metatarsal head, on the mid-foot side of the equinus break between fore-foot and mid-foot. |
| Anterior superior iliac spine | Both left and right |

*# These markers form the minimal marker set.*
**These markers need to be placed asymmetrically to distinguish between left and right in the Vicon plug-in gait lower body model.*

#### Electromyography

Electromyography data will be recorded using Cometa Pico EMG (in Bologna) or Delsys Trigno EMG (in Amsterdam, Kiel, Sydney, and Tel Aviv) at a sample frequency available for each system around 2148 Hz.

Bipolar electrodes will be placed on the leg muscles as listed in Table 5, the first three muscles listed will be measured at each site, whereas the other muscles are optional in this protocol. EMG electrodes will be placed in accordance with the Seniam guidelines for electrode locations (http://www.seniam.org/). Hair will be shaved, and the skin will be cleaned with alcohol prior to attachment of the electrodes. After applying all the electrodes, the EMG signal will be checked by asking the participant to contract the muscles briefly (e.g., abducting the leg a couple of times for gluteus medius muscle, and rotating the ankle a couple of times for the ankle muscles). The power spectrum will be checked for line noise, and precautions will be taken to limit this noise.

*Table 5. Placement of EMG sensors and their reason of interest*

| Muscle | Reason of interest |
|---|---|
| 1. Rectus femoris[#] | Potential role in anteroposterior foot placement stability mechanism |
| 2. Biceps femoris[#] | Potential role in anteroposterior foot placement stability mechanism |
| 3. Gastrocnemius medialis[#] | Potential role in anteroposterior foot placement and push-off stability mechanisms |
| 4. Gluteus medius | Has been shown to contribute to mediolateral gait stability [26,27] |
| 5. Tibialis anterior | Corticomuscular coherence from this muscle has been investigated before in relation to gait [40].<br>Muscle that contributes to mediolateral ankle moment control as a stability mechanism [67]. |
| 6. Peroneus longus | Muscle that contributes to mediolateral ankle moment control as a stability mechanism [67]. |
| 7. Soleus | Muscle that contributes to mediolateral ankle moment control as a stability mechanism [67]. |

*Foot placement control*

To quantify stability-related foot placement control, we will use the foot placement model as previously reported by Wang & Srinivasan (2014), (equation 1).

$$SL = \beta\_pos * CoMpos(i) + \beta\_vel * CoMvel(i) + \varepsilon \qquad [1]$$

### Supplementary material

S1. Questionnaire for healthy participants

S2. Real-world assessment

S3. Analysis plan

S4. Spirit Checklist

S5. Approved ethical protocol
