## Supplementary material for "Steps against the burden of Parkinson’s disease (StepuP): Protocol of a randomized controlled trial elucidating the biomechanical and neurophysiological mechanisms of a speed dependent treadmill training intervention": Approved ethical protocol

#### FULL/LONG TITLE OF THE TRIAL

#### SHORT TRIAL TITLE / ACRONYM

StepuP

#### PROTOCOL VERSION NUMBER AND DATE

Version 1.2; 05. November 2024

SIGNATURE PAGE

The undersigned confirm that the following protocol has been agreed and accepted and that every local Principal Investigator agrees to conduct the trial in compliance with the approved protocol and will adhere to the principles outlined in the Declaration of Helsinki, Good Clinical Practise (GCP) guidelines, and other regulatory requirements.

**Principal Investigator:**

Signature:

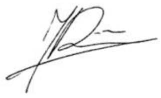

..............................................................................................................

Name: Prof. Jaap van Dieën

Date:
05/11/2024

**Contents**

1. General information ... 6
   i. Key Study Contacts ... 6
2. List of Abbreviations ... 7
   i. Trial Summary ... 8
   ii. Trial Funding ... 10
3. Background ... 10
4. Rationale, objectives, and endpoints ... 11
   i. Rationale ... 11
   ii. Objectives ... 11
   iii. Endpoints ... 11
5. The StepuP trial design ... 12
   i. Study design ... 12
   *ii.* Specific Workpackages ... 12
      A. WP1: Gait improvements after treadmill training ... 12
      B. WP2: Mechanisms underlying gait improvements after treadmill training ... 13
      C. WP3: Daily-life mobility changes after treadmill training ... 13
      D. WP4: Patient engagement and understanding individual responses to treadmill training ... 13
      E. WP5: Management and ECR training ... 13
   iii. Participants ... 13
      A. Inclusion criteria ... 14
      B. Exclusion criteria ... 14
   iv. Participant schedule ... 15
   v. Screening ... 16
   vi. Enrolment ... 16
   vii. Pre-Training (T0) ... 17
      A. Functional assessment ... 17
      B. Home assessment ... 18
      C. Randomization procedures ... 18
   viii. Intervention and control ... 18
      A. General description ... 19
      B. Speed-dependent treadmill single session protocol: details ... 20
      C. Training progression ... 21
      D. Personalized training progression ... 22
   ix. Post-Training (T1) ... 22

### 1. General information

#### i. Key Study Contacts

| | |
|---|---|
| Principal Investigator of the entire project (JPND project No. JPND2022-128) | Professor Jaap van Dieën<br>Vrije Universiteit Amsterdam<br> |
| Funding agencies | Kiel, Germany: Bundesministerium für Bildung und Forschung (www.bmbf.dehttp://www.bmbf.de/)<br>Bologna, Italy: Directorate General for Health Research and Innovation Ministero della Salute (www.salute.gov.it)<br>Sydney, Australia: National Health and Medical Research Council (https://www.nhmrc.gov.au/)<br>Tel Aviv, Israel: Ministry of Health (www.gov.il/en/departments/ministry_of_health)<br>Zurich, Swiss National Science Foundation (https://www.snf.ch/enhttps://www.snf.ch/en)<br>Amsterdam, Netherlands: ZONmw, (https://www.zonmw.nl/) |
| Local Principal Investigators: | Prof. Walter Maetzler, Christian-Albrechts-Universität zu Kiel, Germany<br>Prof. Jeffrey Hausdorff, Tel Aviv Sourasky Medical Center, Israel<br>Dr. Matthew Brodie, University of New South Wales, Sydney, Australia<br>Dr. Fabio La Porta, IRCCS Istituto delle Scienze Neurologiche di Bologna, Italy<br>**Additional partner:**<br>Dr. Navrag Singh, Swiss Federal Institute of Technology, Zürich, Switzerland |
| Data Management Lead | Non-EU:<br>Dr. Martin Ostrowski<br>University of New South Wales, Sydney, Australia<br>EU:<br>Dr. Sjoerd Bruijn<br>Vrije Universiteit Amsterdam, Amsterdam, Netherlands |

#### 2. List of Abbreviations

| | |
|---|---|
| CMC | Cortico-muscular coherence |
| CTT | Color Trail Test |
| CoM | Centre of Mass |
| EEG | Electroencephalography |
| EMG | Electromyography |
| EQ-5D | Euro-Qol (general and validated questionnaire to assess health-related quality of life) |
| FCI | Groll Functional Comorbidity Index |
| FES-I | Falls Efficacy Scale International |
| FI | Frailty Index |
| FACIT | Functional Assessment of Chronic Illness Therapy |
| FOG | Freezing of gait |
| H&Y | Hoehn & Yahr Staging of Parkinson's disease |
| HC | Healthy controls |
| MoCA | Montreal Cognitive Assessment |
| PD | Parkinson’s disease |
| PwPD | Patients with Parkinson’s disease |
| PI | Principal Investigator |
| SDTT | Speed-dependent treadmill training |
| TUG | Timed Up and Go |
| UPDRS | Unified Parkinson’s Disease Rating Scale |
| VAS | Visual Analogue Scale |
| WP | Work Package |
| 2MWT | Two-minute walking test |

#### i. Trial Summary

| | | |
|---|---|---|
| Trial Title | Steps against the burden of Parkinson's Disease | |
| Internal ref. no. (or short title) | StepuP | |
| Trial Design | Multicentre non-pharmacological randomized controlled trial | |
| Participants | Patients with Parkinson's disease (PwPD) recruiting at four different clinical centres and Healthy Controls (HC) recruiting at Amsterdam | |
| Intervention | Speed-dependent treadmill training (i.e, treadmill walking at alternating blocks of different percentages of comfortable walking speed) for 12 sessions with perturbations (sessions between T0 and T1). | |
| Control | Speed-dependent treadmill training without perturbations (12 sessions between T0 and T1). | |
| Maintenance | Both groups receive home-based speed-dependent walk training using a phone app for maintenance of treadmill training benefits between the T1 post-training and T2 follow-up assessments. Includes goal setting towards World Health Organisation physical activity recommendations. | |
| Outcomes | **Measures** | **Domain** |
| | Primary outcome<br>Gait speed (m/sec) | Gait performance |
| | Step width<br>Step width variability<br>Stride time<br>Stride time variability<br>Stride length variability<br>Stride length | Walking |
| | Disease severity (MDS-UPDRS III)<br>Hoehn & Yahr (H&Y)<br>Cognitive function (MoCA & TMT)<br>Depressive symptoms (BDI II)<br>Fear of falling (Short FES-I)<br>Health-related quality of life (EQ-5D)<br>Fatigue (FACIT)<br>Physical capacity (2MWT, TUG, MiniBESTest, Grip strength)<br>Pain and Walking confidence (VAS) | Clinical |

| | | |
|---|---|---|
| | Comorbidities (FCI) | |
| | Local divergence exponent<br>Foot placement model (multiple linear regression) :<br>- Relative explained variance ($R^2$)<br>- Standard deviation of residual (i.e. foot placement error)<br>- Regression coefficients | Kinematics |
| | Beta-band Power in motor areas<br>Cortico-muscular coherence | Neurophysiological |
| | Steps per day<br>Amount and length of uninterrupted walk durations<br>Stride time variability<br>Symmetry | Home assessment |
| | Adverse events<br>Falls and injuries<br>Mobility<br>Fatigue<br>Exercise Adherence<br>Health services use | Weekly Calendars and Safety Monitoring |
| | End-user engagement questionnaires at T1 (regarding treadmill training) and T2 (regarding home-based maintenance walking):<br>- System usability<br>- Physical Activity Enjoyment<br>- Exercise Self Efficacy<br>- Barriers and Enablers to long term use<br>Qualitative interviews using open questions to elicit greater understanding of the individual experiences and barriers and enablers to long term use. | Engagement and Process |
| Planned Trial Period | 24 months | |
| Planned Sample Size | 126 participants with PD<br>21 healthy controls (Amsterdam) | |

#### ii. Trial Funding

This trial is part of a JPND funded consortium awarded in 2022 (Call: "Understanding the mechanisms of non-pharmacological interventions", JPND2022-128)

### 3. Background

Parkinson's disease (PD) affects over 10 million people worldwide and is one of the most prevalent neurodegenerative diseases[1]. It is a multi-system neurodegenerative disorder, which among other symptoms causes severe and disabling motor deficits. One of these deficits, impaired gait, causes falls, a serious complication for people living with PD, even when managed with medication. Approximately 70% of people with PD experience falls each year[2]. The consequences of falls include fractures, hospital admissions, increased caregiver burden, loss of independence, fear of falls, social isolation and early mortality[3]. Falls are cited as one of the worst aspects of PD[4] and unfortunately only few efficacious interventions are available[5]. Non-pharmacological interventions in the form of exercise contribute to slowing disease progression[6] and reduce fall risk through substantial improvements in gait quality, as reflected in increased speed and stride length and reduced gait variability[7,8].

Improving walking ability and preventing falls in people with PD is of critical importance to the world's aging population and health care systems globally. The importance of improving gait and preventing falls for people with PD and their communities is corroborated by our early stakeholder survey. This survey (used in the co-development of this project) included 294 people with PD, 76 family members, 16 clinicians and 29 caregivers. It revealed strong consensus regarding the imperative to develop effective gait training interventions that can improve walking ability, reduce fall risk; and increase quality of life. However, because of gaps in our mechanistic understanding, most exercise prescription fails to address the unmet needs and motor deficits specific to PD. People living with PD walk with short strides and consequently, at a slow speed[18,19], which is associated with poor general health[20]. Their gait pattern is further characterized by a high variability of step characteristics[19], which is associated with falls[21,22].

Fortunately, training can increase stride length and gait speed and reduce variability[7,8]. Treadmill training may provide a response to the unmet needs of people with PD and their community. Generally, exercise interventions targeting gait are effective and more effective than other exercise interventions[11,12], in reducing falls rate and improving gait performance[7-9] and even reducing freezing of gait[10]. Specifically, treadmill training has been shown to be safe and beneficial[9,13,14], and training effects may be enhanced by adding mechanical perturbations and virtual reality[15-17]. Mid-tier treadmills range from €350 to €1,350 and some models can fold away making them practical even in the home situation. Treadmill training is unaffected by inclement weather, can be undertaken several days per week and may be implemented in parallel with pharmacological treatments.

Therefore, the interventions studied here will consist of treadmill training, including the application of mechanical perturbations and virtual reality to destabilize gait and trigger gait adaptations[15-17]. To optimize and personalize treadmill training, it is essential to disentangle how people with PD benefit from treadmill training. To this end, we will relate the positive outcomes of this well-proven non-pharmacological intervention to kinematic, neurophysiological, and psychological changes. We will determine the effects of treadmill training in people with PD at four levels: primary clinical outcomes, feedback control of centre of mass (CoM) movement, neural correlates of feedback control, and mediation of transfer to daily-life gait. In addition, we will explore which factors determine individual outcomes at each of these levels.

### 4. Rationale, objectives, and endpoints

#### i. Rationale

The ***rationale*** of this trial is that speed dependent treadmill training (SDTT) improves gait through improved sensorimotor integration, with changes in cortical activity as neural correlates. Additional benefits of treadmill training can be seen if perturbation or adaptations are added. This is based on the idea, that in addition to the sensorimotor integration the reactive balance in trained as well[18]. Furthermore, it hypothesizes that treadmill training and its effects on gait quality will improve gait self-efficacy, which mediates and/or modifies transfer of training effects to improved daily-life gait.

#### ii. Objectives

Thus, the ***objectives*** of our StepuP project are:

1. To understand the kinematic and neural mechanisms that underlie improvements in gait due to treadmill training with and without mechanically and VR-triggered gait adaptations in people with PD;
2. To assess to what extent improvements in gait due to treadmill training, as measured in the lab, transfer to improvements in daily life mobility;
3. To understand the mechanisms that underpin transfer from improvements in gait to improvements of mobility in daily life in people with PD;
4. To understand for whom treadmill training improves lab-based gait characteristics and for whom it does not, and understand for whom treadmill training improves mobility in daily life and for whom it does not.

Attaining these objectives will provide a better understanding of the successes and failures of treadmill training to improve gait stability and prevent falls in people with PD at an individual level, which in the medium term will allow targeted delivery of such interventions and in the long term will allow personalization of such interventions to improve outcomes for all.

#### iii. Endpoints

Concerning ***the endpoints***, this trial examines the effect of treadmill training with and without perturbations on comfortable overground gait speed, and neural correlates in people with PD. The **primary endpoint** is (change in) gait speed, **secondary endpoints** are divided into three groups (clinical, kinematic and neurophysiological). Clinical measures are used to assess the effect of training on disease symptoms. Kinematic measures are changes from baseline to follow-up, all under controlled conditions (treadmill), and provide insight into gait performance and quality. Neurophysiological measures aim to understand the neural control mechanisms underlying the training effects. On an **exploratory level**, the study aims to assess the training effects on daily-life gait by using wearable devices and assess gait self-efficacy using previously validated questionnaires.

### 5. The StepuP trial design

#### i. Study design

StepuP is a multicenter non-pharmacological randomized controlled trial (RCT) conducted at four clinical centers: Christian-Albrechts-Universität zu Kiel (Kiel), Tel Aviv Sourasky Medical Center (Tel Aviv), University of New South Wales (Sydney), IRCCS Istituto delle Scienze Neurologiche di Bologna (Bologna). 42 PwPD will be enrolled and will be randomized into an intervention group (IG) and a control group (CG) at the clinical sites in Kiel and Sydney. 21 PwPD will be enrolled in Tel Aviv and Bologna and randomized as well into an IG and CG. All Participants will receive a 2-6 week speed-dependent treadmill training (SDTT) aimed at improving gait. Subjects in the IG will receive the same STDD with additional perturbations (SDTT+). Baseline (T0) and follow-up (T1 & T2) visits include descriptive, clinical, kinematic and neurophysiological assessment. A 7-day home monitoring with digital sensors will be performed before T0 and after T2. In Sydney half of the patients in the CG and half of the patients in the IG will have also a second baseline measurement before receiving the SDTT.

#### *ii.* Specific Workpackages

The work within the trial will be broken down into specific work packages (WP), into which the sites mentioned above and other two other sites (Vrije Universiteit Amsterdam, Prof. Dr. JH van Dieën, (Amsterdam) and Swiss Federal Institute of Technology, Dr. Navrag Singh, (Zürich)) contribute. The WP are explained in more detail in the following. Please note that only the clinical centers (Bologna, Kiel, Sydney and Tel Aviv) conduct active recruitment and training, and therefore seek for ethical approval. Figure 1 shows the interaction between WP.

**Figure 1: Overview of WPs of StepuP, and the interaction between them.**

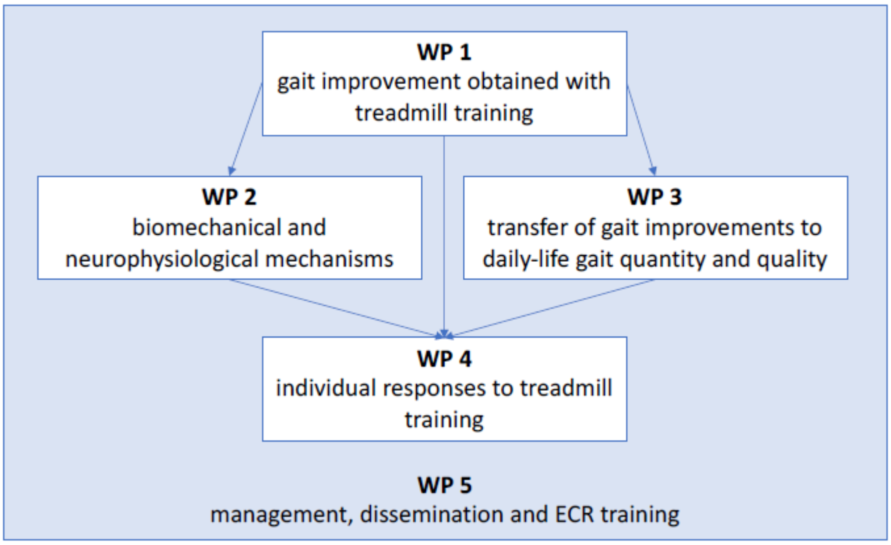

##### A. WP1: Gait improvements after treadmill training

WP1 aims to investigate the gait improvements after treadmill training in patients with Parkinson's disease (PwPD) at four clinical sites (Bologna, Kiel, Sydney, Tel Aviv; WP leader: Kiel). A total of 126 patients will be recruited and undergo training with or without mechanical perturbations or virtual perturbations.

##### B. WP2: Mechanisms underlying gait improvements after treadmill training

This WP (Amsterdam, Bologna, Kiel, Sydney, Tel Aviv; WP leader: Amsterdam) aims to support clinical centers in setting up standardized data collection procedures and to study the factors underlying improvements in primary outcomes related to gait PwPD. The protocol involves assessing lower body kinematics, EMG activity, and EEG data during treadmill training to test the hypothesis that improvements in gait are due to an improved coordination of foot placement to center of mass movement, which is related to better sensorimotor integration. The trial also aims to assess cortical control of locomotion in people with PD and correlate changes in behavioral measures with changes in neural measures. Statistical analyses will focus on correlating changes in primary outcomes with changes in behavioral and neural measures.

##### C. WP3: Daily-life mobility changes after treadmill training

This WP (Bologna, Kiel, Sydney, Tel Aviv, Zürich; WP Leader: Zürich) will analyze the effects of treadmill training on daily-life mobility, focusing on gait quality and physical activity using data from body worn inertial sensors for a week at each time point. The analysis will compare the effects of lab-based training versus real-world training, and examine the relationship between improvements in lab-based gait outcomes and daily-life gait outcomes. The collected data will be used to extract parameters such as gait speed, stride length, and number of steps, and will be analyzed using linear mixed effects models with repeated measures over time. The analysis will also investigate whether gait self-efficacy mediates the effects of lab-based gait improvements on daily-life gait outcomes.

##### D. WP4: Patient engagement and understanding individual responses to treadmill training

The goal of WP4 (Amsterdam, Bologna, Kiel, Sydney, Tel Aviv, Zürich; WP Leader: Sydney), is to combine qualitative interview data with quantitative data from WPs 1-3 to understand why treadmill/enhanced training may improve lab-based gait outcomes and daily-life gait outcomes in some people but not others, and to identify barriers and facilitators to uptake of treadmill training protocols. The team will establish a secure, cloud-based computing environment for GDPR-compliant collection, curation, and analysis of sensitive data. Machine learning will be used to investigate the relative importance of various factors for successful intervention outcomes. The qualitative analysis will provide important insights into barriers and enablers to ongoing treadmill training for developing more personalized and effective therapies to improve walking and prevent falls in people with PD.

##### E. WP5: Management and ECR training

This work package (Amsterdam, Bologna, Kiel, Sydney, Tel Aviv, Zürich; WP Leader: Amsterdam) focuses on project management for the proposed trial on non-pharmacological interventions. The coordinator will oversee project progression and ensure compliance regarding legal and ethical issues, timely action for foreseen or unforeseen risks, and the fulfillment of goals with respect to dissemination, data and code sharing, gender balance, and early-career researcher support. The project will follow data protection and ethical guidelines, use the Brain Imaging Data Structure (BIDS) standard to store motion, EMG, and EEG data, and establish a Github repository for code sharing. The risks of the project are discussed, and working groups will be established to minimize risks. The package also includes support for early-career researchers, including joint supervision, structured research visits, and active involvement in all aspects of the project. ECR workshops on open science will be organized, with training on clinical assessment of patients during the first in-person workshop.

#### iii. Participants

Participants will be enrolled within the study across the four clinical centers on the basis of the following inclusion and exclusion criteria (Table 1):

#### A. Inclusion criteria

1. Diagnosis of PD according to the MDS Criteria[23]
2. Hoehn and Yahr stages I to III;
3. Movement Disorder Society-sponsored version of the Unified Parkinson Disease Rating Scale (MDS-UPDRS) gait sub-score of 1 or more
4. Signed informed consent to participation

#### B. Exclusion criteria

1. Any known general health condition likely to interfere with or to pose a contraindication to physical exercise (e.g., severe heart disease, severe chronic artery disease, orthostatic hypotension according to guidelines, chronic respiratory problems, subjective or objective vertigo, relevant orthopedic and other diseases such as recent lower limbs fractures limiting weight bearing, etc.). For screening and assessment purposes, the 2023 version of the Physical Activity Readiness Questionnaire, extended version (PAR-Q+) will be administered, together with Electronic Physical Activity Readiness Medical Examination (ePARmed-X+) in case of positive screening. Subjects will be excluded if, they are not cleared for engaging in physical activity or are cleared only for medically supervised physical activity.
2. Moderate or severe depression (defined as scores of ≥18 on the Beck Depression Inventory [BDI])
3. Cognitive impairment which may preclude the possibility to provide a fully informed consent to enrollment, as indicated by elements of temporal or spatial disorientation which may be revealed within ordinary conversation or by a confirmed diagnosis of cognitive decline (e.g., Levy-Body Dementia). In case of doubt, the Six-item Cognitive Impairment Test (6CIT, pass cutoff score > 9) will be administered.
4. Linguistic comprehension capacity less than 75% in ordinary conversation due, for instance, to an aphasic impairment or to severe deafness notwithstanding the use of an acoustic aid. In case of doubt, a simple test of linguistic comprehension (token test) will be administered before enrollment.
5. Severe psychiatric comorbidity which may interfere with compliance to the study protocol (e.g., a known diagnosis of personality disorder, documented previous occurrences of delirium and psychomotor agitation).
6. History of or status of substance dependency
7. Implanted Deep Brain Stimulation device

**Table 1: Inclusion and exclusion criteria for participation in the StepuP trial**

| Inclusion Criteria | Exclusion Criteria |
|---|---|
| - PD according to the MDS Criteria<br>- H&Y I-III<br>- MDS-UPDRS gait subscore ≥ 1<br>- Signed informed consent to participation | - Any known general health condition likely to interfere with or to pose a contraindication to non-medically supervised physical exercise.<br>- Moderate or severe depression (BDI-II ≥18)<br>- Cognitive impairment which may preclude the possibility to provide a fully informed consent to enrolment.<br>- Linguistic comprehension capacity less than 75% in ordinary conversation<br>- Severe psychiatric comorbidity which may interfere with compliance to the study protocol<br>- History of or current status of substance dependency<br>- Implanted Deep Brain Stimulation device |

*PD – Parkinson, MDS - Movement Disorder Society, UPDRS Unified Parkinson's Disease Rating Scale, H&Y - Hoehn and Yahr, BDI-II - Beck Depression Inventory-II*

#### iv. Participant schedule

Each participant will undergo a total of four phases subsequent order. An overview can be found in Figure 2. Overview participant's schedule.

**Pre-Training**

The Pre-Training phase including *screening, enrolment, home assessment and functional assessment* (**T0**) will last for two weeks. There should not be more than 14 days between the enrolment and the start of the first training visit. In Sydney, half of the patients who are randomized into the control group and half of those who are randomized into the intervention group, will undertake a second baseline measurement; thus, their *Pre-Training phase* will be extended by 6 weeks. Within these 6 weeks ± 1 week, participants will have undergone the second baseline measurement.

**Training**

The *training* will consist of 12 sessions in total. There can be 0-4 days break between any training session depending of the state of the participant and logistics at each centre.

**Post-Training**

The Post-Training phase including *home assessment and functional assessment* (**T1**) will last for a total of 14 days. There should not be more than 2 days between second functional assessment and the end of the last training visit. Also, the home-assessment should not start later than 5 days after the second functional assessment.

**Follow-Up**

The Follow-Up visit (**T2**) will include a *functional assessment* and take place 12 weeks (±2 weeks) after the completion of the Post-Training phase.

**Figure 2.** Overview participant's schedule.

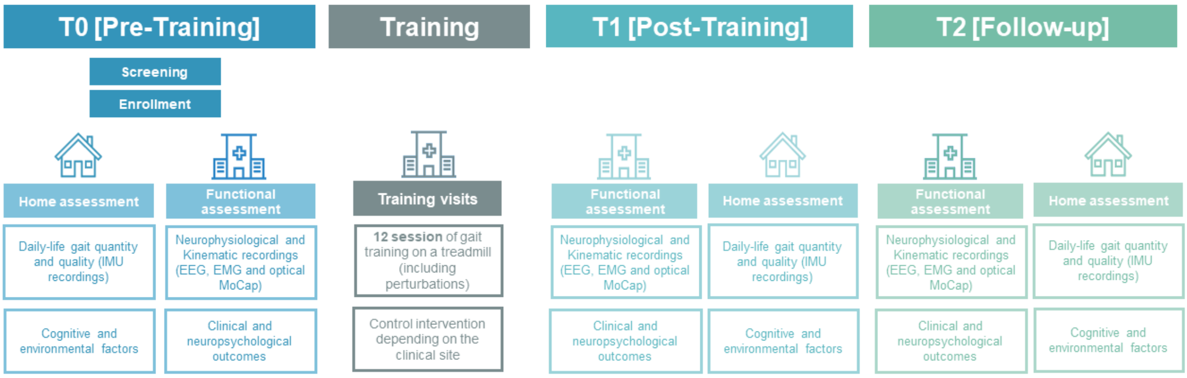

*The Trial consists of a Pre-Training phase including screening, enrolment, home assessment and functional assessment (T0) and a possible second baseline measurement, a Training phase (12 training visits), a Post-Training phase including home assessment and functional assessment (T1) and a Follow-up phase including home assessment and functional assessment (T2).*

#### v. Screening

PwPD: Patients admitted to each of the four clinical centers as in- or out-patients will be invited to participate. Potential candidates can be approached via registries and hospital records (given that they gave priori consent to be contacted for future research) or flyers and local advertisements.

The clinical partner in Kiel can obtain data from clinical routines due to the broad consent statement of a patient which has been hospitalized in the past. In Kiel potential candidates will receive patient information, and the consent form digitally prior to screening.

#### vi. Enrolment

All interested potential participants will attend a screening appointment. This will consist of signing the informed consent, and then reviewing / evaluating the inclusion and exclusion criteria. Prior to signing the informed consent, each person is given sufficient time to consider whether to participate in the study. The researcher must assess that the participant can give consent for themselves. If the assessment does not reveal adequate capacity of the participant, the investigator must usually either exclude the potential participant from the study or seek surrogate consent for their participation. It is made clear to the potential participant, that participation is completely voluntary, and as such they are free to decide to stop participation at any moment without any consequences.

Concerning informed consent, the staff member taking the consent will confirm that the potential participant has read the information sheet before discussing the study further and answering any questions they may have. If the potential participant agrees to participate, they will be asked to sign and date the consent form. This will be witnessed by the researcher taking the consent, who will also sign and date the form. The original consent form is kept in the site file and a copy is given to the participant.

During the preliminary contact, the PI or their delegate (recruiter) will ask the potential candidate if interested in receiving information about the possibility to participate in an non-pharmacological interventional research protocol involving patients with Parkinson's Disease. In case of a positive answer, the recruiter will obtain a consent for communicating the contact information of the patient to the recruiting team. Recruiters will review the inclusion/ exclusion criteria in consultation with a trained neurologist.

The *enrolment* can be completed immediately after *screening*. If this is not completed as a single visit, the visit will be completed within 5 days after the *screening* visit.

#### vii. Pre-Training (T0)

Prior to the *training phase*, participants in all groups across all centers will undergo a standardized *functional assessment*, including measurement of clinical outcomes and a gait analysis during which EEG, EMG and kinematics data will be collected. A video of the participants head will be taken to determine the electrode position. In particular, a computer vision algorithms will detect the position of each electrode in 3D in relation to anatomical. landmarks. This will allow a standardized 2D cartesian projection that could be shared across centers for statistical analysis. Please note that no video files will be shared, only 2D coordinates will be, which do not contain any personalized information[48].

##### A. Functional assessment

*Clinical assessments*

We will assess changes in functional mobility with the standard timed-up-and-go (TUG) test[24,25] and the 2-minute walk test[26], balance and fall risk with the MiniBESTest[27], cognition using the MoCA[32] and CTT[53], and motor symptoms of PD with the MDS-UPDRS[28]. We will use questionnaires to assess effects on gait self-efficacy (modified gait efficacy scale, mGES and falls efficacy scale international, FES-i)[29-31], fatigue (FACIT-F) and health-related quality of life (EQ-5D). After the 2-minute walk test self-rated walking quality and mental effort to walk will be assessed using visual analogue scales.

*Kinematic and neurophysiological measures*

We will assess preferred gait speed using a 20-m walk test (Primary outcome).

Participants start walking on the treadmill for 7 minutes for familiarization. After that, they will then walk at a fixed speed of 1.4 km/h for 5 minutes, to assess the secondary outcomes, step-width and stride-time and the variance of these parameters, from data of at least 100 strides. The fixed 1.4 km/h is attainable for an estimated 95% of patients with moderate PD. However, for those who cannot attain 1.4 km/h at T0, their preferred walking speed T0 will be their fixed speed during all functional assessments. The treadmill assessment will be repeated at their preferred speed. Additionally, during Treadmill walking data from the EEG and EMG will be collected to derive beta-band power in motor areas and calculate cortico-muscular coherence (CMC) between brain and muscle activity.

##### B. Home assessment

A home assessment visit for the period of at least 7 days should be completed. Participants will wear inertial measurement units (IMUs), either a single unit on the lower back or, in a subgroup, a three-sensor system, to continuously record motion data. The sensor on the lower back will be attached using medical tape, while the ankle sensors will be fastened using a self-adjustable band to ensure participant comfort. The sensors can be removed quickly and can be easily repositioned following the instructions provided. The planned number of participants to wear the 3-sensor setup at each site for each interventions follows:

i. 20 people from SDTT (5 each from Sydney, Kiel, Bologna and Tel Aviv)

ii. 16 people from SDTT + VR (8 each from Bologna and Tel Aviv)

iii. 8 people from SDTT + MP (Kiel)

iv. 8 people from SDTT + VR + MP (Sydney)

Participants will be chosen in order of participation until the minimum sample size required is reached.

Participants will be asked to complete a daily diary via their personal smartphone or by paper pencil depending on their preference, including the following information: medication intake, physical activities performed, freezing of gait (FOG) episodes, falls experienced and information on fatigue. Other than the clinical assessment, that is performed during ON medication, the home assessment contains periods of ON, OFF, and transitions between phases, along with various contextual factors that influence mobility[54,55,56]. To accurately evaluate the relationship between improvements in lab-based and daily-life gait outcomes, this diary records the influence of Parkinson's medication and physical activity on mobility during the home assessment and ensures with that comparability between the gait outcomes of T0 and T1. During the functional assessment, participants receive support in personalizing the diary by entering their medication schedule and adding typical physical activities. Over seven days, the diary is completed daily and is expected to take 1–3 minutes per day. Only deviations from the medication schedule need to be noted, while normal intake can simply be checked off for each day. Physical activities, FOG episodes, and falls can be logged by adding abbreviations/X marks in the provided timetable. Participants are encouraged to keep the diary beside their medication box to help prevent late entries, while no daily reminders will be sent to minimize the burden on participants.

##### C. Randomization procedures

To ensure that groups between SDTT and SDTT+ do not differ in Kiel and Sydney, a block wise randomization will be employed. Block size of 5 participants will be used to control for age, and H&Y stage. Randomization is done using a standard algorithm implemented in the Python Package NumPy (see here for details: https://arxiv.org/abs/1805.10941).

#### viii. Intervention and control

#### A. General description

All subjects will receive Speed-Dependent Treadmill Training (SDTT) as a basic intervention. To understand the effects of different kinds of perturbations, each center will do SDTT + additional perturbations. Tel Aviv and Bologna will apply virtual reality (VR) based perturbations, Kiel and Sydney will administer motor perturbations, as shown in Table 2. Intervention and Control groups.

Each intervention program will comprise 12 sessions of 30 minutes each, with a 5-minute warmup and cool down at comfortable overground speed. The comfortable (CS) overground speeds will be determined before each training session using a 20-meter walk test.

**Table 2. Intervention and Control groups.**

| Center | Intervention Group | | Control Group | | Treadmill sessions |
|---|---|---|---|---|---|
| | Treatment | N | Treatment | N | |
| Bologna | SDTT+VR | 11 | SDTT | 10 | 12 |
| Kiel | SDTT+APP | 21 | SDTT | 21 | 12 |
| Sydney | SDTT+VR+APP | 21 | SDTT (2nd baseline) | 21 | 12 |
| Tel Aviv | SDTT+VR | 10 | SDTT | 11 | 12 |

SDTT: Speed-Dependent Treadmill Training; SDTT+VR, SDTT with virtual reality; SDTT+APP, SDTT with anteroposterior perturbations.

The SDTT program is based on proven effective training protocols, and an overhead harness will be used for safety. The physiotherapist will be present but will provide assistance only when occasionally required. The SDTT+ anteroposterior perturbations program will trigger reactive gait adaptations by accelerations and decelerations of one of the belts of a split-belt treadmill, while the SDTT+ multi-directional perturbations will add mediolateral perturbations induced by a sideward translation of the treadmill. If a participant indicates difficulty in completing the training, blocks will be shortened in duration rather than in number and breaks were lengthened. Physiotherapists will only personalize the training progression if needed by omitting or adding a challenge according to the participants' level of exertion[33].

Each SDTT program (control and intervention) will comprise 12 sessions of 30 minutes each. The SDTT+ anteroposterior perturbations (APP) program will trigger reactive gait adaptations by accelerations and decelerations of one of the belts of a split-belt treadmill. The VR SDTT+ is achieved by shifting the visual input horizontally in virtual reality (VR-SDTT+). The SDTT+VR+APP is achieved using a virtual urban street whereby virtual obstacles must be stepped over to avoid induced APP. All perturbation types have been previously applied by us without any adverse events in inactive older adults[33, 34].

After each session, the self-rated walking quality and mental effort visual analogue scales will be recorded and used to track progression and help guide the supervising clinician in tailoring the difficulty of subsequent sessions.

#### B. Speed-dependent treadmill single session protocol: details

*Determining comfortable overground walking speed*

Before the treadmill training, at each first training session of each training week (i.e. the first training session of a set of three training sessions), a single 20-meter walk test will be performed. This test will identify comfortable walking speed (CS).

*Single session protocol on treadmill*

The CS recorded above will be used to set the treadmill speed, in accordance with Figure 3.

Each training session consists of three walking training trials at speeds V1, V2 and V3, intermitted with recovery walking trials. V1 will always be 80% of the comfortable walking speed, whereas V2 and V3 will be determined as the percentage of CS that progresses across training sessions (see "Training progression" for more detail). Prior to the first training trial, between each training trial and at the end of the last trial participants will walk at 70% of CS, as warmup/recovery walking trials. The warmup/recovery trials will last 2.5 minutes, unless longer breaks are necessary for the participant to tolerate the protocol. During walking, participants will be encouraged to use reciprocal arm swing (unless they are holding the handrails in the first minute of a new speed interval, see above).

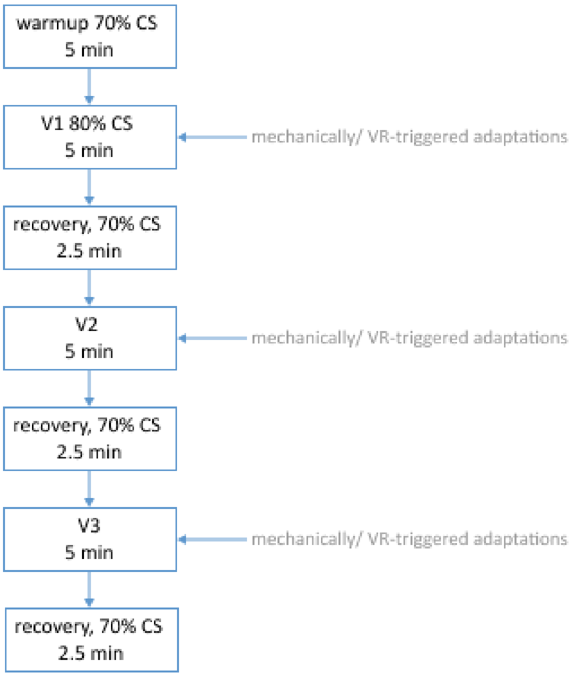

**Figure 3.** Overview of a single session of the SDTT

*Methods to ensure safety*

During the treadmill walking participants will wear an overhead safety harness which does not support the participant's weight during walking, but will prevent them from falling. Treadmill railings will be used only if needed and only during the first minute of speed intervals to adjust to the speed demands.

A physiotherapist/clinician will be present to monitor the participant fatigue and gait pattern but will not give any physical assistance during the walking.

If possible, the physiotherapist will aim for completion of the training protocol (i.e., the total treadmill walking duration will remain the same) for the participant but will ensure sufficient breaks are inserted to maintain safety for the participant. During these resting breaks the participant will be seated and is allowed to drink water and or eat some snacks. Frequency and duration of these breaks will be reported.

#### C. Training progression

The challenge of the training is expected to increase with the performance of the participant, as every first training session of the week the comfortable overground walking speed is re-assessed. So within a week participants will walk based on the same comfortable walking speed, but across weeks the speed may increase.

In addition, the %CS is progressed for V2 and V3 and the duration of the walking training trials (V1,V2,V3) will be lengthened over training sessions (Table 3). Progression of the VR challenge will be automatic based on the participant's performance, and the perturbation challenge will be increased across sessions as well.

*Table 3. Progression of training speed and duration*

| *Training session #* | *CS of session #* | *V1* | *V2* | *V3* | *Training trial duration progression* | *Max training trial duration (full progression)* |
|---|---|---|---|---|---|---|
| *1* | *1* | *80% CS* | *85% CS* | *90% CS* | *5 min** | *5 min* |
| *2* | *1* | *80% CS* | *85% CS* | *90% CS* | *+30 sec* | *5 min 30 sec* |
| *3* | *1* | *80% CS* | *90% CS* | *95% CS* | *=* | *5 min 30 sec* |
| *4* | *4* | *80% CS* | *90% CS* | *95% CS* | *+30 sec* | *6 min* |
| *5* | *4* | *80% CS* | *95% CS* | *100% CS* | *=* | *6 min* |
| *6* | *4* | *80% CS* | *95% CS* | *100% CS* | *+30 sec* | *6 min 30 sec* |
| *7* | *7* | *80% CS* | *100% CS* | *105% CS* | *=* | *6 min 30 sec* |
| *8* | *7* | *80% CS* | *100% CS* | *105% CS* | *+ 30 sec* | *7 min* |
| *9* | *7* | *80% CS* | *100% CS* | *110% CS* | *=* | *7 min* |
| *10* | *10* | *80% CS* | *100% CS* | *105% CS* | *+30 sec* | *7 min 30 sec* |
| *11* | *10* | *80% CS* | *100% CS* | *105% CS* | *=* | *7 min 30 sec* |

| | | | | | | |
|---|---|---|---|---|---|---|
| *12* | *10* | *80% CS* | *100% CS* | *110% CS* | *+ 30 sec* | *8 min* |

** Or maximum tolerated duration*

##### D. Personalized training progression

Physiotherapists will personalize the training progression according to the participants' level of exertion. This personalization of the training progression will be recorded. A record will be kept regarding V1, V2 and V3, total duration of walking, and duration of rests at each session. In personalizing the participant's progression, the guidelines below will be followed.

*Personalized progression of speed*

Speeds are only increased by 5% at V2 and V3 if the previous maximum attained speed was tolerated. If after starting any walking training trial (V1/V2/V3) the speed is not tolerated, the speed will be lowered 5% to the previously tolerated walking speed before the bout is resumed.

*Personalized progression of duration*

If the participant tolerates the speed but not the duration of the walking training period, the duration of V2 and V3 will be shortened (-30 seconds) to the previously tolerated walking duration during the training trials. In addition, they are given a longer recovery walking period at 70% CS (+30 seconds) and on top of this a seated resting break can be included if needed.

*Personalized progression of VR*

An automatic progress mode in the system allows VR progression to be personalized, based on the subjects performance.

#### ix. Post-Training (T1)

After each *training phase*, participants in all groups across all centers will undergo another *functional assessment*, including measurement of clinical outcomes and a gait analysis during which EEG, EMG and kinematics data will be collected. As done at T0 a video of the participants head will be taken to determine the electrode position.

##### A. Functional assessment

*Clinical assessments*

We will assess changes in functional mobility with the standard timed-up-and-go (TUG) test[24,25] and the 2-minute walk test[26], balance and fall risk with the MiniBESTest[27], and motor symptoms of PD with the MDS-UPDRS[28]. We will use questionnaires to assess effects on gait self-efficacy (modified gait efficacy scale, mGES and falls efficacy scale international, FES-i) [29-31], fatigue (FACIT-F) and health-related quality of life (EQ-5D).

*Kinematic and neurophysiological measures*

We will assess preferred gait speed and maximal gait speed using a 20-m walk test (Primary outcome).

Participants start walking on the treadmill for 7 minutes for familiarization. After that, they will then walk at their preferred speed for 5 minutes, to assess the secondary outcomes, step-width and stride-time and the variance of these parameters, from data of at least 100 strides. The fixed 1.4 km/h is attainable for an estimated 95% of patients with moderate PD. However, for those who cannot attain 1.4 km/h at T0, their preferred walking speed T0 will be their fixed speed during all functional assessments. The treadmill assessment will be repeated at their preferred speed. Additionally, during Treadmill walking data from the EEG and EMG will be collected to derive beta-band power in motor areas and calculate cortico-muscular coherence (CMC) between brain and muscle activity.

#### B. Home assessment

A home assessment visit for the period of at least 7 days should be completed. Participants will wear inertial measurement units (IMUs), either a single unit on the lower back or, in a subgroup, a three-sensor system, to continuously record motion data. The sensor on the lower back will be attached using medical tape, while the ankle sensors will be fastened using a self-adjustable band to ensure participant comfort. The sensors can be removed quickly and can be easily repositioned following the instructions provided. The planned number of participants to wear the 3-sensor setup at each site for each interventions follows:

i. 20 people from SDTT (5 each from Sydney, Kiel, Bologna and Tel Aviv)

ii. 16 people from SDTT + VR (8 each from Bologna and Tel Aviv)

iii. 8 people from SDTT + MP (Kiel)

iv. 8 people from SDTT + VR + MP (Sydney)

Participants will be chosen in order of participation until the minimum sample size required is reached.

Participants will be asked to complete a daily diary via their personal smartphone or by paper pencil depending on their preference, including the following information: medication intake, physical activities performed, freezing of gait (FOG) episodes, falls experienced and information on fatigue. Other than the clinical assessment, that is performed during ON medication, the home assessment contains periods of ON, OFF, and transitions between phases, along with various contextual factors that influence mobility[54,55,56]. To accurately evaluate the relationship between improvements in lab-based and daily-life gait outcomes, this diary records the influence of Parkinson's medication and physical activity on mobility during the home assessment and ensures with that comparability between the gait outcomes of T0 and T1. During the functional assessment, participants receive support in personalizing the diary by entering their medication schedule and adding typical physical activities. Over seven days, the diary is completed daily and is expected to take 1–3 minutes per day. Only deviations from the medication schedule need to be noted, while normal intake can simply be checked off for each day. Physical activities, FOG episodes, and falls can be logged by adding abbreviations/X marks in the provided timetable. Participants are encouraged to keep the diary beside their medication box to help prevent late entries, while no daily reminders will be sent to minimize the burden on participants.

##### C. Qualitative interviews

Online questionnaires will be used to assess:

- System usability
- Physical Activity Enjoyment
- Exercise Self Efficacy
- Barriers and Enablers to long term use

Furthermore, space for participants to provide feedback and suggestions for future improvements will be used to record all participants' experiences of the treadmill training.

#### x. Maintenance Period (T1 to T2)

##### A. Home-based walking

All participants will be offered a home-based speed dependent walk training intervention. This intervention is an App based training for gait adaptability and allows users to set their own training time and pace. It delivers a rhythmic metronomic beat for three different walking speeds, designed to trigger movement and encourage better walking patterns. The Walking Tall app can be installed on iOS and Android devices and does not store any personalized data. During the maintenance period, participants will be supported using the app to progress towards World Health Organisation recommendations for moderate intensity exercise through a personalised goal setting session.

##### B. Weekly Calendars

Weekly online calendars will be used to assess:

- Physical activity both related to the study and other planned activities
- Falls adverse events
- Falls and injuries
- Mobility
- Fatigue
- Exercise Adherence both related to the study and other planned activities
- Health services use

Reported falls will be followed up with a phone call to assess severity and ensure ongoing participant safety.

#### xi. Follow-up assessment (T2)

After each 12 weeks (±2 weeks), participants in all groups across all centers will undergo another *functional assessment*, including measurement of clinical outcomes and a gait analysis during which EEG, EMG and kinematics data will be collected. As done at T0 and T1 a video scan of the participants head will be taken to determine the electrode position.

#### A. Functional assessment

*Clinical assessments*

We will assess changes in functional mobility with the standard timed-up-and-go (TUG) test[24,25] and the 2-minute walk test[26], balance and fall risk with the MiniBESTest[27], and motor symptoms of PD with the MDS-UPDRS[28], cognition using the MoCA[32] and CTT. We will use questionnaires to assess effects on gait self-efficacy (modified gait efficacy scale, mGES and falls efficacy scale international, FES-i) [29-31] fatigue (FACIT-F) and health-related quality of life (EQ-5D). After the 2-minute walk test self-rated walking quality and mental effort to walk will be assessed using visual analogue scales.

*Kinematic and neurophysiological measures*

We will assess preferred gait speed and maximal gait speed using a 20-m walk test (Primary outcome).

Participants start walking on the treadmill for 7 minutes for familiarization. After that, they will then walk at their preferred speed for 5 minutes, to assess the secondary outcomes, step-width and stride-time and the variance of these parameters, from data of at least 100 strides. The fixed 1.4 km/h is attainable for an estimated 95% of patients with moderate PD. However, for those who cannot attain 1.4 km/h at T0, their preferred walking speed T0 will be their fixed speed during all functional assessments. The treadmill assessment will be repeated at their preferred speed. Additionally, during Treadmill walking data from the EEG and EMG will be collected to derive beta-band power in motor areas and calculate cortico-muscular coherence (CMC) between brain and muscle activity.

*Qualitative interviews*

To meet the JPND grant guidelines, which includes a focus on recording end-user experiences participants, in-person or remote/telephone structured interview at T2 will be performed on enrolled participants. Answers to the open-ended questions (as detailed in the interview guide) will be analysed using qualitative methods to understand the participants' experiences, enablers and barriers to intervention success. The planned number of participants necessary to be interviewed at each site for each arm in order to achieve saturation is as follows:

- 20 people from SDTT (5 each from Sydney, Keil, Bologna and Tel Aviv)
- 10 people from SDTT + VR (5 each from Bologna and Tel Aviv)
- 10 people from SDTT + MP (Kiel)
- 10 people from SDTT + VR + MP (Sydney)

These will provide data for qualitative analyses of both cultural context and intervention type differences. Interim qualitative analyses will be performed at monthly intervals in order to check whether saturation is achieved.

Online questionnaires will be used to assess:

- System usability
- Physical Activity Enjoyment

- Exercise Self Efficacy
- Barriers and Enablers to long term use

Furthermore, space for participants to provide feedback and suggestions for future improvements will be used to record all participants' experiences of the home-based walk training.

##### B. Home assessment

A home assessment visit for the period of at least 7 days should be completed. Participants will wear inertial measurement units (IMUs), either a single unit on the lower back or, in a subgroup, a three-sensor system, to continuously record motion data. The sensor on the lower back will be attached using medical tape, while the ankle sensors will be fastened using a self-adjustable band to ensure participant comfort. The sensors can be removed quickly and can be easily repositioned following the instructions provided. The planned number of participants to wear the 3-sensor setup at each site for each interventions follows:

i. 20 people from SDTT (5 each from Sydney, Kiel, Bologna and Tel Aviv)

ii. 16 people from SDTT + VR (8 each from Bologna and Tel Aviv)

iii. 8 people from SDTT + MP (Kiel)

iv. 8 people from SDTT + VR + MP (Sydney)

Participants will be chosen in order of participation until the minimum sample size required is reached.

Participants will be asked to complete a daily diary via their personal smartphone or by paper pencil depending on their preference, including the following information: medication intake, physical activities performed, freezing of gait (FOG) episodes, falls experienced and information on fatigue. Other than the clinical assessment, that is performed during ON medication, the home assessment contains periods of ON, OFF, and transitions between phases, along with various contextual factors that influence mobility[54,55,56]. To accurately evaluate the relationship between improvements in lab-based and daily-life gait outcomes, this diary records the influence of Parkinson's medication and physical activity on mobility during the home assessment and ensures with that comparability between the gait outcomes of T0 and T1. During the functional assessment, participants receive support in personalizing the diary by entering their medication schedule and adding typical physical activities. Over seven days, the diary is completed daily and is expected to take 1–3 minutes per day. Only deviations from the medication schedule need to be noted, while normal intake can simply be checked off for each day. Physical activities, FOG episodes, and falls can be logged by adding abbreviations/X marks in the provided timetable. Participants are encouraged to keep the diary beside their medication box to help prevent late entries, while no daily reminders will be sent to minimize the burden on participants.

#### xii. Outcome variables

The following outcome variables will be collected. For an easier overview clinical assessment has been collectively referred to outcomes A-E.

#### A. Descriptive Measures

A1. General descriptive measures: Year of birth, gender, height, weight, shoe size, education, employment, living arrangement, current physical or other therapies, smoking history, alcohol consumption at T0.
A2. Treatment acceptance and satisfaction to the training and overall protocol will be assessed to T1 and T2.

#### B. Clinical outcome measures

B1. Fall events (occurrence and frequency) and fall related injuries: The number of falls and whether the falls were injurious will be recorded. Twelve-month retrospective during T0 and during the duration of the trial[2].
B2. Fracture history: Number and type of fracture sustained will be recorded. Twelve-month retrospective during T0.
B3. Medication: Current medication will be recorded at each visit.
B4. Euro-Qol (EQ-5D): The EQ-5D measures quality of life[42]. It consists of two components; health state description and evaluation. This will be completed on T0, T1 and T2.
B5. Frailty Index (FI): The FI measures frailty five different criteria (shrinking, low physical endurance/energy, low physical activity, weakness and slow walking speed). This will be completed at T0, T1 and T2.
B6. Functional Assessment of Chronic Illness Therapy (FACIT) Fatigue scale[43]: The FACIT Fatigue Scale measures fatigue during usual daily activities over the past week. This will be completed at T0, T1 and T2.
B7. Visual Analogue Scale (VAS)[38]: The VAS will be used to determine the pain during rest and walking and the self-assessed quality of walking. This will be completed at T0, T1 and T2.
B8. Groll Functional Comorbidity Index (FCI)[39]: This is a self-administered test with physical function as the outcome. This will be completed at T0.

#### C. Physical measures

C1. Gait speed: Preferred overground gait speed (primary outcome) and maximal overground gait speed (secondary outcome) using a 20-m walk test[45] will be derived and expressed in meters per second. The test will be conducted at T0, T1 and T2.
C2. Use of mobility aids: The use of commonly used walking aids (indoor and outdoor) will be recorded. This will be recorded at T0 and T1.
C3. Timed Up and Go (TUG)[24,25]: The TUG is a common clinical measure used to assess mobility, balance, and walking ability in older adults. This will be completed at T0, T1 and T2.
C4. Two-minute walking test (2MWT): The 2MWT is used to measure functional exercise capacity. The distance in meters covered in 6 minutes is recorded. This will be completed at T0, T1 and T2.
C5. Mini-BESTest: The Mini-BESTtest[27] is used to assess an individual's balance and stability on a series of functional tasks at T0, T1 and T2.
C6. Modified Gait Efficacy Scale (mGES)[30,31] is used to assess walking confidence under challenging everyday circumstances and will be completed at T0, T1 and T2.
C7. Hand grip strength[40]: This will determine the maximal grip strength of the participant. The test will be completed at T0.

#### D. Neuropsychological measures

D1. Short Falls Efficacy Scale International (Short FES-i): The short FES-i[29] is a measure the level of concern about falling during social and physical activities inside and outside the home. This will be completed at T0, T1 and T2.

D2. Montreal Cognitive Assessment (MoCA): The MoCA[32] is a measure of cognitive impairment. It assesses different cognitive domains: attention and concentration, executive functions, memory, language, visuo-constructional skills, conceptual thinking, calculations, and orientation. This will be completed at T0 and T2.

D3. Color Trail Test (CTT): Color Trail Test [44] is a neuropsychological assessment that assesses a person's cognitive flexibility, visual scanning and executive function. It involves connecting a series of numbered circles, alternating between numbers and letters in ascending order as quickly as possible, and provides insight into attention, mental flexibility and processing speed. This will be completed at T0 and T2.

##### E. Parkinson-Specific Assessments

E1. Movement Disorder Society Unified Parkinson's Disease Rating Scale (MDS-UPDRS): The MDS-UPDRS[28] describes disease progression. It is separated into four different domains including cognitive function, behaviour and mood, activities of daily living (ADL) and motor examination. This will be completed at T0, T1 and T2.

E2. New Freezing of Gait Questionnaire (NFOGQ): The NFOGQ[41] is a tool to detect and evaluate the impact and severity of freezing of gait. This will be completed at T0, T1 and T2.

##### F. Kinematics and Neurophysiological measures

F1. Kinematics: 3D motion capture data will be recorded at T0, T1 and T2.

F2. Electroencephalography (EEG): EEG records an electrical signal from the surface of the scalp and can be used to describe brain activity. This will be recorded at T0, T1 and T2.

F3. Electromyography (EMG): Surface EMG records the electrical activity of superficial muscles through electrodes placed on the skin, and it is commonly used in clinical and research settings to assess muscle function, fatigue, and activation patterns. This will be recorded at T0, T1 and T2.

##### G. Home assessment

G1. The steps per day will be recorded at T0 and T1.
G2. The amount and length of uninterrupted walk durations will be determined (T0 & T1)
G3. The stride time variability will be derived at T0 and T1.
G4. Symmetry of gait will be determined at T0 and T1.
G5. FOG episodes, Medication and Physical activity will be recorded by self-reports (T0 and T1)
G6. Weekly Calendars will be used to assess Physical activity, Falls and Injurious Falls.

##### H. Engagement and Process Outcomes (T1 and T2)

H1. Acceptability and satisfaction with the intervention will be assessed with the 10-item System Usability Scale[49], Physical Activity Enjoyment Scale (PACES)[50] and questions developed specifically for the components of this study.
H2. Changes in attitudes towards physical activity will be assessed with the Exercise Self-Efficacy Scale (ESES)[51]
H3. Exit interviews will be conducted at to explore the barriers and enablers to long-term use and reasons for ceased involvement (if applicable) using pre-prepared, open-ended questions according to qualitative interviews methods .

#### xiii. Safety and risk management procedures

Clinical treadmill training with a harness is considered a safe and effective intervention for patients with Parkinson's disease (PD). The use of a harness provides additional support and reduces the risk of falls during treadmill training, allowing patients to focus on their gait and balance training without fear of injury. In this respect, during the intervention, the risk of adverse events is not increased compared to what may be expected during regular daily life activities. Clinical treadmill training with a harness is a safe and effective intervention that can improve the physical and functional abilities of patients with PD.

Falls, falls-related injuries, myalgia and fatigue are considered potentially trial-associated adverse events (AE) and will thus be monitored for the duration of the trial. We will assess the number of falls and the number of injurious falls during the previous 12-months at T0, throughout the duration of the trial and the weeks preceding T2.

An AE form should be completed and returned to the local Principal Investigator within 24 hours of the site's awareness of the event. This form must be reviewed and signed off by the PI will determine the seriousness and causality in conjunction with the trial procedures.

#### xiv. Bias minimization procedures

To reduce bias, evaluations of participants will be done in a blinded manner (i.e. without knowledge of whether they belong to a SDTT group or a SDTT+ group, or whether the participant will take part in a second baseline measurement). Any data collection at T0 will be done before the assignment of a patient. The clinical assessment at T1 and T2 will be run by personal, which will not know the group the participant was assigned to. Quantitative outcome parameters (e.g. EEG parameters or gait outcomes) are not prone to be bias.

#### xv. Specific administrative procedures

**Withdrawal from the study**

All participants remain free to withdraw at any time from the trial without giving reasons and without prejudicing their further treatment and must be provided with a contact point where he/she may obtain further information about the trial. Participants will be informed, that upon request all data collected up to the point of withdrawal will be permanently deleted.

**Reimbursement policy**

Participants can receive reimbursement of reasonable expenses, according to local guidelines. Travel expenses may include own vehicle, public transport or taxi.

#### xvi. Data management procedures

To make data management GDPR compliant, we will use the VU research drive, hosted by the VU in Amsterdam, Netherlands to provide a central access point to protocols, standards and workflows, and to streamline the creation and curation of a central data asset based on the FAIR (Findable, Accessible Interoperable and Reusable) principles. To ensure high standards of data processing, the widely accepted Brain Imaging Data Structure (BIDS) standard to store the motion, EMG and EEG data will be followed [36,37]. A Github repository will be established to facilitate code sharing and a communication channel will be established to facilitate day to day interactions between partners. The channel will specifically support early-career researchers, to enhance knowledge transfer, skills sharing and mentoring across the network. Code developed in WP2 and WP3 for data analysis will be fully shared in the consortium and will be made publicly available upon publishing results, via the MJFF and OpenNeuro.

Participant data will be collected in a coded, de-identified manner. Data will be stored on a RedCap server hosted in the EU in a standardized form across the StepuP consortium. This data will be organized following BIDS standards[36, 37]. Only pseudonymized data will be shared between partners using European servers.

All recruitment sites will keep original records of all signed consent forms, trial key codes, and any other paper forms or samples that are collected at source, under secure conditions at the site or origin until the trial has been completed and the database has been locked. After this point, these documents can be purged or archived for a further period, depending on local requirements.

Direct access will be granted to authorized representatives from the PI, host institution and the regulatory authorities.

Clinical sites will be responsible to archiving all trial documents for a period of time that is in keeping with institutional or national guidelines that pertain to that site. Destruction of documentation should be notified to the PI.

##### xvii. End of trial

The end of trial will be defined date of the last visit/data item of the last participant undergoing the trial.

##### xviii. Dissemination policy

Dissemination of project results is crucially important to reach a long-lasting impact. StepuP has multiple measures in place to maximise dissemination of the results emphasising an Open Access policy and is committed to this goal by employing several strategies. Emphasizing an Open Access policy, the project actively participates in conferences to engage with relevant stakeholders, furthering the reach and understanding of its findings. In a notable move towards inclusivity, StepuP has also planned to invite patients to present the final results, aligning the research more closely with those directly affected. Moreover, the project recognizes the importance of media exposure and plans to utilise local channels (e.g. newspapers) for sharing information with the broader community.

#### 6. Statistics and analysis plan

##### *i.* Sample size calculation

The sample size was calculated using the SampSize application[1]on for the clinical primary objective (i.e., comfortable gait speed overground, measured in m/sec from a 20 meters walking test[45]), with a significance level (alpha) of 0.05 and power (1-beta) of 0.80, with an allocation ratio of 1:1 in parallel groups. From the available literature, it is known that standard treadmill (SDTT) may improve comfortable overground gait speed up to 0.1 m/sec (from a baseline of 1.26 m/sec)[46] in comparison to standard overground walking and that SDTT with perturbations (SDTT+) may further increase confortable overground gait speed up to +0.08 m/sec in comparison to SDTT[47]. Thus, we will assume that SDDT+ is superior to SDTT at improving comfortable overground gait speed based on an expected average difference in gait speed between the two groups of +0.08 m/sec with a SD of 0.08[46,47] Under this assumption, 17 subjects per group are expected to be enrolled, rising to 20.4 considering a maximum drop-out rate of 20%. This would lead to 41 subjects per trial, rounded to 42 to allow for groups of equal size.

Overall, 126 patients at all clinical sites will in enrolled over a period of 24 months. The recruitment totals for each site is outlined in Table 2. Given the time frame of the trial, recruitment will be monitored on a monthly basis. Should any site encounter recruitment difficulties, other sites have the capacity to recruit additional participants for each cohort as well as to recruit additional cohorts if necessary.

##### ii. Analysis plans and datasets

The statistical analysis of the outcomes will comprise the definition of analysis sets and statistical analysis (including prioritization of outcomes).

[1] https://app.sampsize.org.uk

The main aim is to determine the mechanisms underlying effects on gait performance in clinical tests and in daily-life of SDTT. This entails an analysis of pooled data (excluding the healthy control participants). Nested within this design, several randomized comparisons will be made to assess the added value of the SDTT+ components over SDTT. Initially a placebo comparison was planned but given the proven effect of SDTT this was deemed unethical. To interpret the any effects changes in the absence of a placebo control group it is first of all important to determine the minimal meaningful change for the outcome parameters. In addition, a comparison between participants with and without PD will be done to support interpretation of effects in the PD group. These analysis to this end are described as analysis 1a and 1b. Subsequently the nested randomized comparisons are described as levels 2a to 2c. Finally, the analysis to test hypothesis on the causality of changes in gait performance will be described as analyses 3a and 3b.

#### iii. Descriptive analysis

Main characteristics of patients, including detailed description of walking capacity, will be done by number and percentage for categorical variables, mean and standard deviation for continuous variables with normal distribution, and median and percentiles 25$^{th}$-75$^{th}$ for continuous variables with non-normal distribution.

#### iv. Analysis of reliability (1a)

**Population:**

From the populations measured in Sydney 21 participants chosen at random will participate in an additional measurement set 14 days prior to start of the intervention (T-1). This sample is kept small for pragmatic reasons. First, it is not feasible to apply this additional measurement in all participants at all centers. Second, in case a systematic effect of repeated testing is found, this may need to be statistically corrected for or the sub-sample of individual participants with this additional baseline may need to omitted in the pooled analyses (3a and 3b).

**Dependent variables:**

*primary clinical outcomes:*

- preferred gait speed (20 m walk test)
- stride length (at preferred speed)
- stride length (at 1.4 km/h)

*secondary clinical outcomes:*

- stride length variability (at preferred speed and at 1.4 km/h)
- stride time (at preferred speed and at 1.4 km/h)
- stride time variability (at preferred speed and at 1.4 km/h)
- step width (at preferred speed and at 1.4 km/h)
- step width variability (at preferred speed and at 1.4 km/h)
- falls (diary)
- timed-up-and-go (TUG) test
- 2-minute walk test
- MiniBESTest
- MOCA
- MDS-UPDRS, part III
- mGES

*feedback control variables (all at 1.4 km/h):*

- $R^2$ (feedback model relating foot placement to centre of mass movement)
- RMS error (feedback model relating foot placement to centre of mass movement)
- Position gain (feedback model relating foot placement to centre of mass movement)

- Velocity gain (feedback model relating foot placement to centre of mass movement)

*neural correlates (T1 all at 1.4 km/h):*

- EEG beta power in(pre-)motor areas during walking at preferred speed
- EEG-EMG coherence during walking at preferred speed

**Analysis technique:**

Bland-Altman analyses will be performed to test for a systematic effect of repeated measurements (paired t-tests) and to establish the limits of agreement for the outcome variables. The limits of agreement thus established will provide the basis for evaluating any effect sizes found in analyses 2a-2c.

#### v. Case-control study (1b)

**Population:**

21 (min. 17 for analysis) PD patients (T0) randomly selected from total sample

21 (min. 17 for analysis) healthy controls age- and sex-matched to PD group

**Dependent variables:**

*primary clinical outcomes:*

- preferred gait speed (20 m walk test)
- stride length (at preferred speed and at 1.4 km/h)

*secondary clinical outcomes:*

- stride length variability (at preferred speed and at 1.4 km/h)
- stride time (at preferred speed and at 1.4 km/h)
- stride time variability (at preferred speed and at 1.4 km/h)
- step width (at preferred speed and at 1.4 km/h)
- step width variability (at preferred speed and at 1.4 km/h)

*feedback control variables (at 1.4 km/h):*

- $R^2$ (feedback model relating foot placement to centre of mass movement)
- RMS error (feedback model relating foot placement to centre of mass movement)
- Position gain (feedback model relating foot placement to centre of mass movement)
- Velocity gain (feedback model relating foot placement to centre of mass movement)

*neural correlates (at 1.4 km/h):*

- beta power during walking at preferred speed
- EEG-EMG coherence during walking at preferred speed

**Analysis technique:**

Initially, independent t-tests and for EEG data cluster-based permutation tests time locked to the gait cycle will be used to compare patients to controls. The presence of and direction of differences between groups will facilitate interpretation for any effects found in analyses 2a - 2c. For this no corrections for multiple comparisons will be made. However, to present overall differences between groups we will reduce dimensionality of the set of primary and secondary clinical outcomes by means of PCA and use stepwise logistic regression on PC-scores to assess between group differences. For feedback control variables and neural correlates we will separately correct for multiple comparisons.

##### vi. SDTT vs. SDTT+ (2a)

Added value of additional component (SDTT+) to Speed Dependent Treadmill Training (SD Analysis technique.

**Dependent variables:**

*primary clinical outcomes:*

- preferred gait speed (20 m walk test)
- stride length (at preferred speed)
- stride length (at 1.4 km/h)

*secondary clinical outcomes (T1):*

- stride length variability (at preferred speed and at 1.4 km/h)
- stride time (at preferred speed and at 1.4 km/h)
- stride time variability (at preferred speed and at 1.4 km/h)
- step width (at preferred speed and at 1.4 km/h)
- step width variability (at preferred speed and at 1.4 km/h)
- falls (diary)
- timed-up-and-go (TUG) test
- 2-minute walk test
- MiniBESTest
- MOCA
- MDS-UPDRS, part III
- mGES

*secondary clinical outcomes (T2):*

- steps per day
- uninterrupted walk durations
- stride time variability
- symmetry

*feedback control variables (T1 all at 1.4 km/h):*

- $R^2$ (feedback model relating foot placement to centre of mass movement)
- RMS error (feedback model relating foot placement to centre of mass movement)
- Position gain (feedback model relating foot placement to centre of mass movement)
- Velocity gain (feedback model relating foot placement to centre of mass movement)

*neural correlates (T1 all at 1.4 km/h):*

- EEG beta power in(pre-)motor areas during walking at preferred speed
- EEG-EMG coherence during walking at preferred speed

Linear mixed effect models with the interaction of treatment and time in the model and a random intercept for participant nested within center (Twisk et al., 2018), controlling for relevant potential confounders (e.g., age, H&Y stage). For the EEG data, we will use cluster-based permutation tests time locked to the gait cycle.TT)

##### vii. Independent RCTs SDTT vs SDTT+ (3a)

**Independent variables:**

time (T0,T1,T2) x group (SDTT, SDTT+)

**Dependent variables:**

*primary clinical outcomes:*

- preferred gait speed (20 m walk test)
- stride length (at preferred speed)
- stride length (at 1.4 km/h)

*secondary clinical outcomes (T1):*

- stride length variability (at preferred speed and at 1.4 km/h)
- stride time (at preferred speed and at 1.4 km/h)
- stride time variability (at preferred speed and at 1.4 km/h)
- step width (at preferred speed and at 1.4 km/h)
- step width variability (at preferred speed and at 1.4 km/h)
- falls
- timed-up-and-go (TUG) test
- 2-minute walk test
- MiniBESTest
- MOCA
- MDS-UPDRS, part III
- mGES

*secondary clinical outcomes (T2):*

- steps per day
- uninterrupted walk durations
- stride time variability
- symmetry

*feedback control variables (T1 all at 1.4 km/h):*

- $R^2$ (feedback model relating foot placement to centre of mass movement)
- RMS error (feedback model relating foot placement to centre of mass movement)
- Position gain (feedback model relating foot placement to centre of mass movement)
- Velocity gain (feedback model relating foot placement to centre of mass movement)

*neural correlates (T1 all at 1.4 km/h):*

- EEG beta power in(pre-)motor areas during walking at preferred speed
- EEG-EMG coherence during walking at preferred speed

**Analysis technique:**

Linear mixed effect models with the interaction of treatment and time in the model and a random intercept for participant (Twisk et al., 2018), controlling for relevant potential confounders (e.g., age, H&Y stage). For the EEG data, we will use cluster-based permutation tests time locked to the gait cycle.

#### viii. Mechanisms underlying changes in gait performance (3b)

**Population:** 126 PD patients

Associations between clinical outcomes and feedback control

**Dependent variables:**

*primary clinical outcomes (T1-T0 / T2-T0):*

- Δ preferred gait speed (20 m walk test)
- Δ stride length (at preferred speed and at 1.4 km/h)

*secondary clinical outcomes (T1-T0 / T2-T0):*

- Δ stride length variability (at preferred speed and at 1.4 km/h)

- Δ stride time (at preferred speed and at 1.4 km/h)
- Δ stride time variability (at preferred speed and at 1.4 km/h)
- Δ step width (at preferred speed and at 1.4 km/h)
- Δ step width variability (at preferred speed and at 1.4 km/h)

**Independent variables:**

*feedback control variables (T1-T0 / T2-T0, at 1.4 km/h):*

- Δ $R^2$ (feedback model relating foot placement to centre of mass movement)
- Δ RMS error (feedback model relating foot placement to centre of mass movement)
- Δ Position gain (feedback model relating foot placement to centre of mass movement)
- Δ Velocity gain (feedback model relating foot placement to centre of mass movement)

*neural correlates (T1-T0 / T2-T0, at 1.4 km/h):*

- Δ beta power during walking at preferred speed
- Δ EEG-EMG coherence during walking at preferred speed

Associations between feedback control variables and neurological correlates

**Dependent variables:**

feedback control variables (T1-T0 / T2-T0, at 1.4 km/h):

- Δ $R^2$ (feedback model relating foot placement to centre of mass movement)
- Δ RMS error (feedback model relating foot placement to centre of mass movement)
- Δ Position gain (feedback model relating foot placement to centre of mass movement)
- Δ Velocity gain (feedback model relating foot placement to centre of mass movement)

**Independent variables:**

neural correlates (T1-T0 / T2-T0, at 1.4 km/h):

- Δ beta power during walking at preferred speed
- Δ EEG-EMG coherence during walking at preferred speed

**Analysis technique:**

We will correlate changes in primary outcomes (e.g., walking speed) with changes in behavioural (foot placement coordination) and neural (EEG beta band power, cortico-muscular coherence) outcomes. Multilevel models will be used, to account for data collected at different sites. For outcomes T2-T0, potential predictors of clinical outcomes will include independent variables with a time lag (T1-T0) and without time lag (T2-T0).

##### ix. Daily life Mobility (3c)

During the home assessment (T0 and T1), participants will wear miniature inertial measurement units (IMUs) (23 x 32.5 x 7.6 mm) for seven days to capture their daily life mobility. The IMUs will be affixed to the body using medical adhesive tapes (e.g., lower back) and/or straps (e.g., on the ankle). The sensors include an onboard data logger that is suitable for collecting longitudinal movement data. The feasibility of collecting such data has already been demonstrated through the UK Biobank study (https://www.ukbiobank.ac.uk/), which included nearly 100,000 participants, and in clinical populations such as Parkinson's disease in the ongoing MobiliseD project (Micó-Amigo et al., 2023).

The collected IMU movement data will undergo post-processing to extract walking bouts and their respective lengths. Following the definition by Micó-Amigo et al. (2023), a walking bout is defined as 'a walking sequence containing at least two consecutive strides of both feet.' To evaluate daily life gait quantity, parameters such as steps/strides per day and the number of turns will be derived. Quality of gait will be assessed based on the extracted walking bouts, including the calculation of parameters such as stride length, stride variability, and gait symmetry.

Sample size was calculates based on the desired precision of the foot placement control estimate[52]. SD and error margin are estimated from pilot data on foot placement control in PD:

$$N = \left(\frac{z * sd}{error}\right)^2$$

$$N = \left(\frac{1.96 * 0.25}{0.07}\right)^2 = 49$$

**Population:** 126 PD patients

Associations between daily life gait and gait performance

**Dependent variables:**

*secondary clinical outcomes (T1-T0):*

- Δ steps per day
- Δ uninterrupted walk durations
- Δ stride time variability
- Δ symmetry

**Independent variables:**

*primary clinical outcomes (T1-T0):*

- Δ preferred gait speed (20 m walk test)
- Δ stride length (at preferred speed)

*secondary clinical outcomes (T1-T0):*

- Δ stride length variability (at preferred speed)
- Δ stride time (at preferred speed)
- Δ stride time variability (at preferred speed)
- Δ step width (at preferred speed)
- Δ step width variability (at preferred speed)

*feedback control variables (T1-T0):*

- Δ $R^2$ (feedback model relating foot placement to centre of mass movement)
- Δ RMS error (feedback model relating foot placement to centre of mass movement)
- Δ Position gain (feedback model relating foot placement to centre of mass movement)
- Δ Velocity gain (feedback model relating foot placement to centre of mass movement)

*gait efficacy (T1-T0):*

- Δ mGES

**Analysis technique:**

We will correlate changes in daily-life outcomes with changes in lab-based gait performance outcomes. Multilevel models will be used to account for data collected at different sites. We will perform mediation analysis to assess the role of gait efficacy. In addition, in an exploratory analysis we will use machine learning to generate prediction models of the dependent variables listed here based on the independent variables listed and patient characteristics such as age, disease duration, and disease severity.

##### x. Factors influencing individual treatment success (3c)

**Aims:**

This WP will focus on the qualitative (interview) data combined with quantitative data from previous analyses to determine:

(i) why treadmill/enhanced training may improve lab-based gait outcomes in some but not other people; and

(ii) why lab-based gait outcomes may transfer to improved daily-life gait in some but not other people;

We aim to provide a holistic understanding of the mechanisms underlying the transfer of changes in clinic-based gait outcomes to changes in real-world mobility outcomes, their retention and the barriers and enablers to wide spread treatment uptake and long-term acceptance.

**Population:** 126 PD patients

**Analysis technique:**

We will focus on understanding the differences between individuals who do and do not improve on the lab-based tests, and individuals who do and do not improve in their daily-life related outcomes. We will use machine learning to investigate, for example, the relative importance to successful intervention of: (i) baseline demographics, education, sex, age, and socioeconomic status; (ii) experiences and preferences of persons with PD; (iii) clinical gait and balance outcomes; (iv) changes in lab-based gait and mechanistic outcomes; and (v) social and psychological factors for individual outcomes in daily-life, such as mobility, quality of life. This is critical towards developing more personalized and long-term effective therapies that improve walking and prevent falls in people with PD.

### 7. Ethical and regulatory considerations

#### i. Regulatory Compliance

The trial will be conducted according to the declaration of Helsinki, Good Clinical Practise (GCP) standards and The European Code of Conduct for Research Integrity. The protocol will be submitted to ethical approval from the local ethical boards. Once approved, any changes to the protocol will be submitted as an emendament for ethical approval and, once approved, the relevant stakeholders (funding authorities, trial registries) will be informed. The aim of this trial is to gain a better understanding of the underlying mechanisms that contribute to the symptoms of PD, and to identify potential treatment options that may be effective in reducing those symptoms. We therefore argue that according to the Medical Devices Act, paragraph §§19ff, this constitutes a trial in the context of basic research, and is therefore not a clinical investigation under the Medical Devices Act. All devices used in this trial are CE marked and FDA-certified. Special attention is given to the compliance and compatibility of EU regulations regarding local regulatory requirements outside EU (e.g. in Australia, Israel and Switzerland).

#### ii. Data protection and patient confidentiality

Data will only be shared between sites in coded and de-identified form. The key linking participants' names will be securely kept by the main researchers of each measurement site.

#### iii. Access to the final trial dataset

All members of the StepuP team will have access to the coded and de-identified final trial dataset. After publication the coded and de-identified data will be open access published on OpenNeuro for further use of dataset beyond the goals of this project.
